# Vision Loss in Neurocysticercosis: Clinical Features, Diagnostic Approaches, and Treatment Outcomes: a systematic review of case report and case series

**DOI:** 10.1101/2024.07.23.24310899

**Authors:** Ravindra Kumar Garg, Pragati Garg, Vimal Kumar Paliwal, Shweta Pandey

## Abstract

**Background:** Neurocysticercosis is a parasitic infection caused by *Taenia solium* larvae, leading to various neurological symptoms, including vision loss. This systematic review critically analyzes cases of vision loss associated with neurocysticercosis to evaluate diverse etiologies and assess the visual prognosis.

**Methods:** This review follows the PRISMA guidelines and is registered with the International Prospective Register of Systematic Reviews (registration number: CRD42024556278). Reports involving human subjects with vision loss due to neurocysticercosis were included. Databases searched included PubMed, Scopus, Embase, and Google Scholar.

**Results:** A total of 148 records, encompassing 168 patients, were included. The mean age was 27.3 years, with 41 percent females and 58.3 percent males. The majority of cases were from Asia. The duration of illness varied, with most cases spanning one month to six months. Common symptoms included headache or orbital pain. Vision loss was predominantly unilateral. Imaging abnormalities included multiple cystic brain lesions, enhancing brain lesions, and calcified lesions. The most affected structures were the intravitreal and retinal regions, followed by the anterior chamber, orbital apex, and optic nerve. Anticysticercal drugs were the main treatment, with 60.12 percent showing improvement.

**Conclusions:** Vision loss in neurocysticercosis is primarily due to the involvement of the intravitreal and retinal regions, with frequent occurrence of multiple cystic brain lesions.

Anticysticercal treatment leads to significant improvement in many cases, although surgical intervention is often required for intravitreal or retinal cysts. The prognosis varies, with the majority of patients had improved vision, though some experience severe outcomes such as eye enucleation.

## Introduction

Neurocysticercosis, caused by the larval stage of the pork tapeworm *Taenia solium*, remains a significant global health challenge, particularly in regions such as Latin America, sub-Saharan Africa, and Southeast Asia. This parasitic infection primarily targets the central nervous system and is recognized as a leading cause of acquired epilepsy worldwide. Transmission occurs through the ingestion of eggs present in contaminated food or water, or via direct fecal-oral contact. The clinical spectrum of neurocysticercosis includes seizures, headaches, visual disturbances, and various neurological deficits. Diagnostic modalities encompass neuroimaging techniques, including magnetic resonance imaging and computed tomography scans, supplemented by serological tests to confirm the diagnosis. The therapeutic approach involves the use of antiparasitic agents like albendazole and praziquantel, corticosteroids, antiepileptic drugs, and, in certain cases, surgical intervention.^**1,2**^

Vision loss in neurocysticercosis is a severe and debilitating complication arising from multiple pathophysiological mechanisms. These include direct ocular involvement by the cysts, elevated intracranial pressure, and inflammatory responses impacting the optic nerves. The clinical presentation of vision impairment can vary from mild visual disturbances to complete blindness and is frequently associated with other neurological symptoms such as seizures and headaches.^**3-5**^

Most of the information about vision loss in neurocysticercosis is derived from case reports and case series. Therefore, in this review, we will critically analyze these cases to evaluate the diverse etiologies contributing to vision loss in neurocysticercosis and to assess the visual prognosis associated with this condition.

## Methods

This systematic review follows the guidelines from the PRISMA statement, which gives rules for how to do reviews and meta-analyses. Our review is registered with PROSPERO (registration number: CRD42024556278).^**6**^ We did not need approval from the Institutional Ethics Committee because we did not involve any human subjects.

### Inclusion and Exclusion Criteria

This systematic review focused on studies related to vision loss in neurocysticercosis. We included studies available in any language that involved human subjects and provided information on the clinical features, diagnostic approaches, and treatment outcomes for vision loss associated with neurocysticercosis. We excluded reports that did not specifically address vision loss in neurocysticercosis, as well as animal studies, in vitro studies, conference abstracts, and review articles. We used revised diagnostic criteria provided by Del Brutto and colleagues for establishing the diagnosis of neurocysticercosis.^**7**^

### Literature Search Strategy

The databases of PubMed, Scopus, Embase, and Google Scholar were systematically searched for relevant literature. In Google Scholar, the search extended through the first 50 pages of results. No language restrictions were applied, and non-English articles were translated using Google Translate. The search terms included (“vision loss” OR “visual impairment” OR “blindness” OR “visual acuity” OR “visual field” OR “optic neuropathy” OR “ocular manifestation” OR “eye disease” OR “sight loss”) AND (“neurocysticercosis” OR “NCC” OR “*Teania solium*” OR “cysticercosis” OR “parasitic brain infection”). The most recent search was done on 10 June, 2024.

### Data extraction

Two independent researchers (RK and VP) screened the titles and abstracts of search results to identify studies meeting the criteria for vision loss in neurocysticercosis. Full texts of potentially relevant studies were thoroughly assessed. Any discrepancies in study inclusion were resolved through discussion or consultation with a third reviewer (PG). In cases of differing opinions, consensus was reached through discussion.

EndNote 21 software (Clarivate, Philadelphia, PA, USA) was used to manage duplicate entries, with any discrepancies or issues resolved through consensus. A PRISMA flow diagram was prepared, detailing the number of records obtained and assessed at each stage, facilitated by the EndNote 21 tool.

### Quality assessment

Each case was evaluated based on four key criteria: selection, ascertainment, causality, and reporting, as outlined by Murad MH et al.^**8**^ According to the standards set by Della Gatta et al., a case report meeting all these criteria was deemed “good quality.” A report meeting three criteria was rated “fair quality,” while those meeting only one or two criteria were considered “poor quality.”^**9**^

### Data Analysis

Demographic details, clinical features, neuroimaging of the central nervous system, anatomical location, types and numbers of lesions in the brain or ocular structures, diagnostic procedures, final diagnoses, treatments, and patient outcomes were recorded in a Microsoft Word file. The consolidated data was then presented in tables, displaying the corresponding numbers and percentages.

## Results

Our review encompassed 148 records, accounting for a total of 168 patients (Supplementary Item 1). Figure 1 displays the PRISMA flowchart used in our systematic review. Out of the 168 cases analyzed, 142 (85%) were determined to be of good or fair quality, whereas 26 (15%) cases were categorized as low quality (Supplementary Item 2). However, none the low quality cases was excluded from the analysis. The PRISMA checklist is available as Supplementary Item 3.

**Figure-1.**
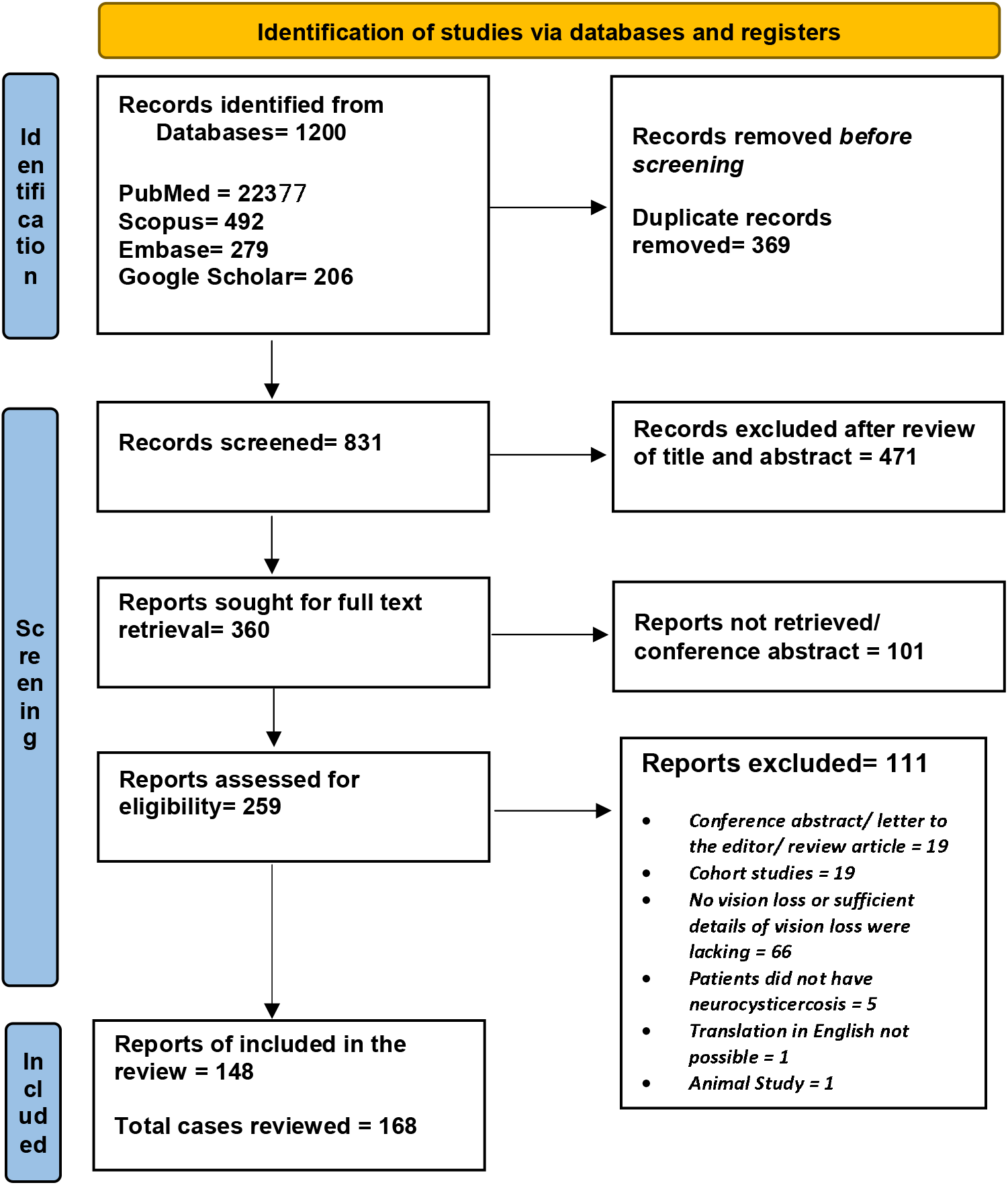
PRISMA flow diagram of the study depicts the procedure of selecting articles for the systematic review.

In this systematic review, the mean age of reviewed cases was 27.3 years (median 25; range 1-73 years). There were a total of 69 (41%) females and 98 (58.3%) males. Among 148 reviewed records, Asia accounted for 67.6% of the total, with India contributing 56.8% and Nepal 4%. Europe had 5.4%, Africa 4%, North America 18.2%, and South America 4.7%. (Table-1)

**Table-1:**
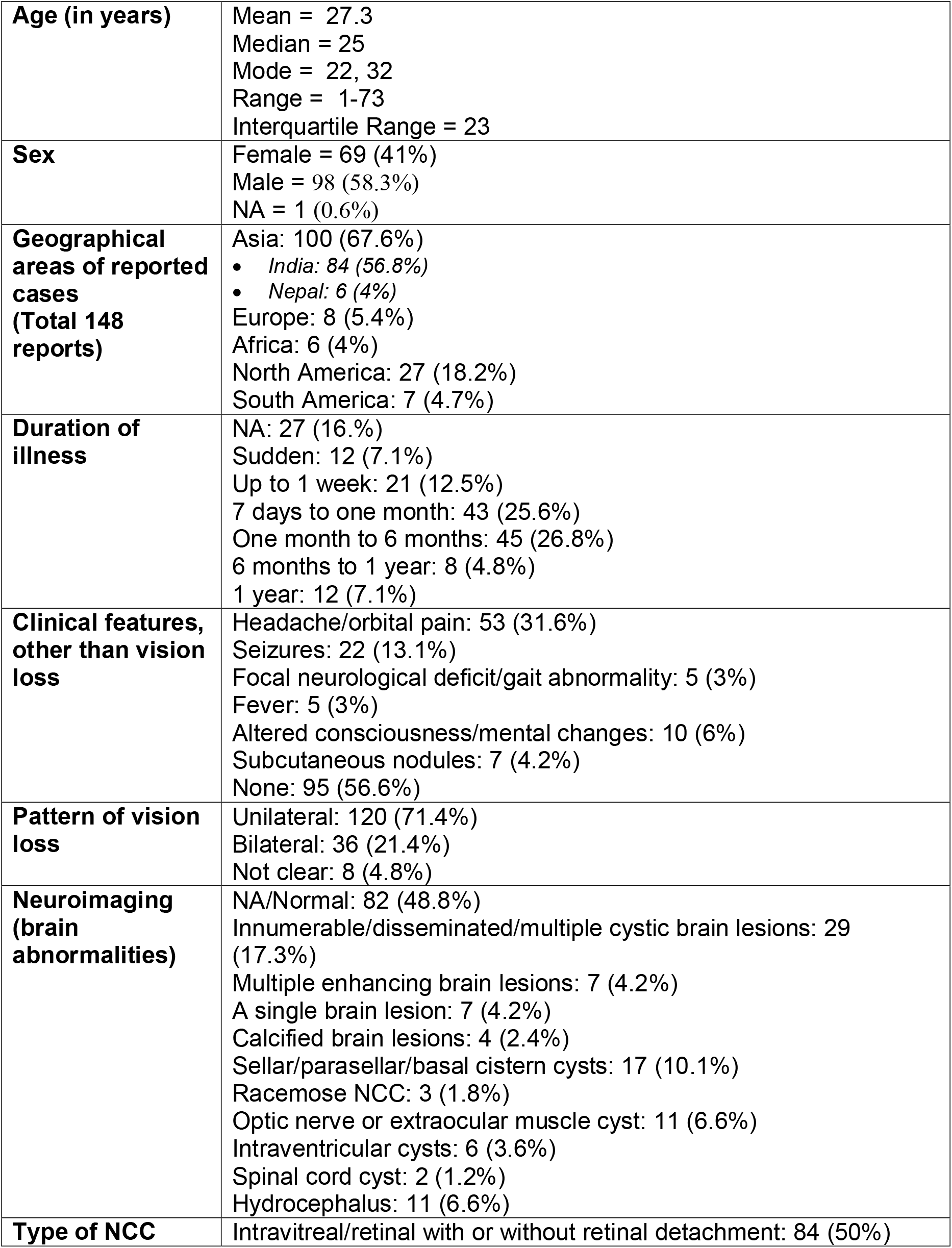
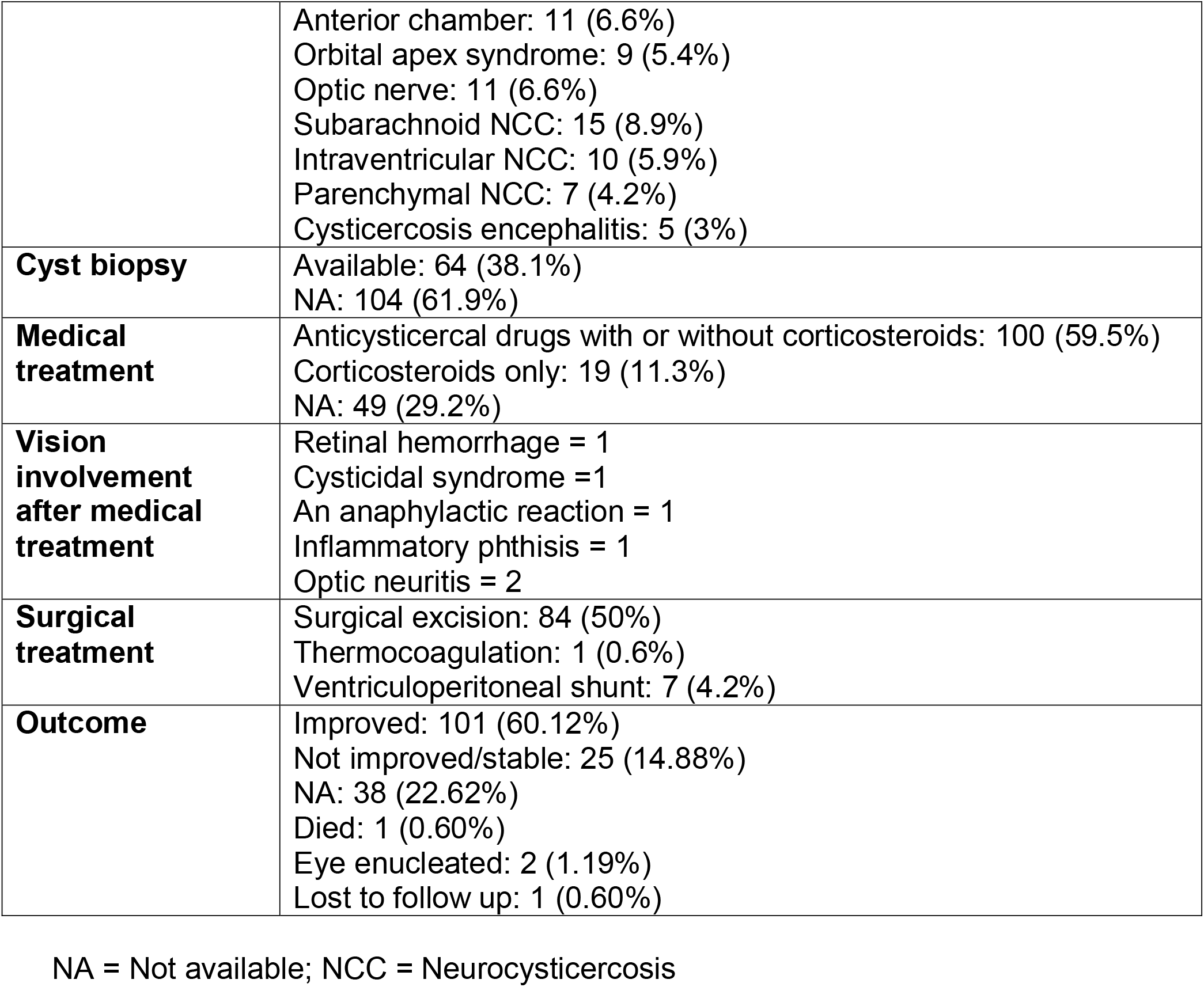
A summary of the clinical features, neuroimaging characteristics, and treatment outcomes in patients with neurocysticercosis who experienced vision loss (n = 168)

|  |  |
| --- | --- |
| **Age (in years)** | Mean = 27.3<br>Median = 25<br>Mode = 22, 32<br>Range = 1-73<br>Interquartile Range = 23 |
| **Sex** | Female = 69 (41%)<br>Male = 98 (58.3%)<br>NA = 1 (0.6%) |
| **Geographical areas of reported cases (Total 148 reports)** | Asia: 100 (67.6%)<br>India: 84 (56.8%)<br>Nepal: 6 (4%)<br>Europe: 8 (5.4%)<br>Africa: 6 (4%)<br>North America: 27 (18.2%)<br>South America: 7 (4.7%) |
| **Duration of illness** | NA: 27 (16.%)<br>Sudden: 12 (7.1%)<br>Up to 1 week: 21 (12.5%)<br>7 days to one month: 43 (25.6%)<br>One month to 6 months: 45 (26.8%)<br>6 months to 1 year: 8 (4.8%)<br>1 year: 12 (7.1%) |
| **Clinical features, other than vision loss** | Headache/orbital pain: 53 (31.6%)<br>Seizures: 22 (13.1%)<br>Focal neurological deficit/gait abnormality: 5 (3%)<br>Fever: 5 (3%)<br>Altered consciousness/mental changes: 10 (6%)<br>Subcutaneous nodules: 7 (4.2%)<br>None: 95 (56.6%) |
| **Pattern of vision loss** | Unilateral: 120 (71.4%)<br>Bilateral: 36 (21.4%)<br>Not clear: 8 (4.8%) |
| **Neuroimaging (brain abnormalities)** | NA/Normal: 82 (48.8%)<br>Innumerable/disseminated/multiple cystic brain lesions: 29 (17.3%)<br>Multiple enhancing brain lesions: 7 (4.2%)<br>A single brain lesion: 7 (4.2%)<br>Calcified brain lesions: 4 (2.4%)<br>Sellar/parasellar/basal cistern cysts: 17 (10.1%)<br>Racemose NCC: 3 (1.8%)<br>Optic nerve or extraocular muscle cyst: 11 (6.6%)<br>Intraventricular cysts: 6 (3.6%)<br>Spinal cord cyst: 2 (1.2%)<br>Hydrocephalus: 11 (6.6%) |
| **Type of NCC** | Intravitreal/retinal with or without retinal detachment: 84 (50%) |
|  | Anterior chamber: 11 (6.6%)<br>Orbital apex syndrome: 9 (5.4%)<br>Optic nerve: 11 (6.6%)<br>Subarachnoid NCC: 15 (8.9%)<br>Intraventricular NCC: 10 (5.9%)<br>Parenchymal NCC: 7 (4.2%)<br>Cysticercosis encephalitis: 5 (3%) |
| **Cyst biopsy** | Available: 64 (38.1%)<br>NA: 104 (61.9%) |
| **Medical treatment** | Anticysticercal drugs with or without corticosteroids: 100 (59.5%)<br>Corticosteroids only: 19 (11.3%)<br>NA: 49 (29.2%) |
| **Vision involvement after medical treatment** | Retinal hemorrhage = 1<br>Cysticidal syndrome = 1<br>An anaphylactic reaction = 1<br>Inflammatory phthisis = 1<br>Optic neuritis = 2 |
| **Surgical treatment** | Surgical excision: 84 (50%)<br>Thermocoagulation: 1 (0.6%)<br>Ventriculoperitoneal shunt: 7 (4.2%) |
| **Outcome** | Improved: 101 (60.12%)<br>Not improved/stable: 25 (14.88%)<br>NA: 38 (22.62%)<br>Died: 1 (0.60%)<br>Eye enucleated: 2 (1.19%)<br>Lost to follow up: 1 (0.60%) |

NA = Not available; NCC = Neurocysticercosis

The duration of illness was quite variable. The highest percentage (26.8%) of occurrences spanned one month to six months, followed by 25.6% occurring between seven days to one month. Events lasting up to one week accounted for 12.5%, and those classified as sudden made up 7.1%. Instances lasting exactly one year were also 7.1%, while events between six months to one year accounted for 4.8%.(Table-1)

Among a cohort of 168 cases, headache or orbital pain was present in 31.6% of cases, followed by seizures in 13.1%. Focal neurological deficits or gait abnormalities and fever each accounted for 3%. Altered consciousness or mental changes were observed in 6%, and subcutaneous nodules in 4.2%. Notably, 56.6% of cases reported no symptoms other than vision loss. The pattern of vision loss showed that 71.4% of cases were unilateral, 21.4% were bilateral, and in the rest, it was not clearly defined. (Table-1)

The data described various neuroimaging abnormalities in 168 cases. In 48.8% of cases, neuroimaging was either normal or not available. Innumerable, disseminated, or multiple cystic brain lesions were recorded in 17.3% of cases, while multiple enhancing brain lesions and single brain lesions each accounted for 4.2%. Calcified brain lesions were seen in 2.4%, and sellar/parasellar/basal cistern cysts in 10.1%. Racemose neurocysticercosis occurred in 1.8%, optic nerve or extraocular muscle cysts in 6.6%, intraventricular cysts in 3.6%, and spinal cord cysts in 1.2%.

Hydrocephalus was present in 6.6% of cases. (Table-1)

In 168 cases of neurocysticercosis with vision loss, the anatomical structures involved included intravitreal and retinal regions, with or without retinal detachment, in 50% of cases. The anterior chamber was affected in 6.6%, and the orbital apex syndrome was present in 5.4%. The optic nerve was involved in 6.6%, while subarachnoid neurocysticercosis was seen in 8.9%. Intraventricular neurocysticercosis occurred in 5.9%, parenchymal neurocysticercosis in 4.2%, and cysticercosis encephalitis in 3% of the cases. (Table-1)

In the medical treatment of 168 patients with neurocysticercosis and vision loss, 59.5% received anticysticercal drugs with or without corticosteroids, 11.3% were treated with corticosteroids only, and in 29.2% medical treatment was not offered. Only 6 patients reported visual side effects following anticysticercal therapy. In half of the cases, the cysts were surgically removed. In these cases, cysts were either in the intravitreal/retinal region or were in the anterior chamber. (Table-1)

In the prognosis of 168 neurocysticercosis patients with vision loss, 60.12% improved, 14.88% remained stable or did not improve, 22.62% were not applicable, 0.60% died and 1.19% had an eye enucleated. (Table-1)

## Discussion

In this review of 168 neurocysticercosis patients with vision loss, significant findings included various brain imaging abnormalities such as multiple cystic brain lesions, enhancing brain lesions, and calcified brain lesions. Approximately, 17% of patients either had number of cysts “too numerous to count” or disseminated neurocysticercosis. Key affected anatomical structures included the intravitreal and retinal regions, with involvement of the anterior chamber, orbital apex, optic nerve, subarachnoid, intraventricular, and parenchymal areas. The most common symptoms were headache or orbital pain. Anticysticercal drugs were the primary treatment, leading to improvement in many patients. Patients with Intravetreous/ retinal cysts required surgical intervention. The prognosis varied, with a majority showing improvement while a few experienced severe outcomes like eye enucleation.

We noted that ocular neurocysticercosis, particularly Intravetreous neurocysticercosis, was the most frequent cause of vision loss. Cases of cysticercosis involving the anterior chamber, vitreous cavity, and subretinal space were categorized as intraocular segment cysticercosis. It is speculated that cysticercosis reaches the eye through the choroidal circulation, which has vessels with a higher flow rate. In the eye, the macula is a preferred site due to its rich blood supply. Cysticerci can migrate from the macula to the vitreous cavity, potentially causing retinal detachment or scarring. Once in the vitreous cavity, the larvae develop into cysticerci, triggering an immune response that leads to vitreous inflammation and various visual impairments, including floaters, blurred vision, and reduced visual acuity, with severe cases resulting in complications like vitreous hemorrhage, retinal detachment, or endophthalmitis.^**10**^ High-resolution ultrasonography is crucial for diagnosing and monitoring orbital and adnexal cysticercosis. B-scan ocular ultrasonography reveals a well-defined cyst with a hyperechoic scolex, providing clear visualization.^**11**^ Treatment generally involves the surgical removal of living parasites and many cases are treated with anticysticercal drugs, albendazole, along with corticosteroids.^**12-14**^ The fear that administering systemic anticysticercal drugs to patients with ocular cysticercosis may trigger inflammatory reaction associated with severe visual impairments. Our review noted that only in 6 out of 168 cases heightened inflammatory reaction with worsening of visual impairments were noted.

Another important cause of vision loss in neurocysticercosis is because of the presence of cysts in sellar/ parasellar region. A rare form of neurocysticercosis involves the development of cysts in the basal subarachnoid region, known as racemose neurocysticercosis. Racemose neurocysticercosis often manifests as meningeal, intraventricular, or subarachnoid (cisternal) forms. Subarachnoid form of neurocysticercosis leads to raised intracranial pressure, widespread meningitis, and CSF obstruction, causing hydrocephalus, vasculitis, and strangulation of optic nerves and optic chiasma, which results in vision loss. ^**15**^ In an cohort of Chang and Keane examined 23 cases of vision loss due to neurocysticercosis and noted optic neuropathy, optic chiasmal damage, and retrochiasmal lesions as primary causes.

Optic neuropathy, often linked to papilledema and hydrocephalus, was a significant factor. Chiasmal damage resulted from cyst-related inflammation and compression. Retrochiasmal lesions were due to large parenchymal cysts or vasculitic cerebral infarction.^**16**^

In our review, about 17% of neurocysticercosis cases with vision loss had the disseminated form of the disease. Extracranial dissemination most frequently affects the skin, subcutaneous tissues, and muscles. In a systematic review, among 222 cases of disseminated neurocysticercosis, vision loss was reported in 46 (21%) cases. Vision impairment in disseminated neurocysticercosis occurs due to cysticercal encephalitis or the presence of cysts in the optic nerve or retina.^**17**^

Cysticercal encephalitis, characterized by extensive brain inflammation from numerous inflamed cysticerci, typically contraindicates the use of cysticidal drugs, as rapid parasite elimination can exacerbate inflammation, increase intracranial pressure, and risk trans-tentorial herniation. High-dose corticosteroids, particularly dexamethasone, have proven effective in managing this condition by reducing inflammation and associated risks. In cysticercal encephalitis, vision is impacted due to increased intracranial pressure.^**18**^

The key limitations of the review is a publication bias that limit generalizability. Additionally, the variable quality of included reports restrict the ability to draw definitive conclusions. The heterogeneous nature of the cases, variations in diagnostic criteria, and differences in treatment approaches further complicate the analysis.

In conclusion, vision loss in neurocysticercosis mainly arises from intravitreal and retinal involvement, often alongside multiple brain cysts. Anticysticercal treatment significantly improves many cases, but surgical intervention is frequently needed for intravitreal or retinal cysts. While most patients experience improved vision, some may face severe outcomes, including eye enucleation.

## Supporting information

Supplementary item-1

Supplementary item-2

Supplementary item-3

## Data Availability

All data produced in the present work are contained in the manuscript

## The ethical statement

No human or animal subjects were involved so ethical clearance was not taken.

