## Supplementary item-1 for "Vision Loss in Neurocysticercosis: Clinical Features, Diagnostic Approaches, and Treatment Outcomes: a systematic review of case report and case series"

**Supplementary table 1: Details of clinical features, neuroimaging characteristics and treatment outcomes in patients of neurocysticercosis with vision loss (n=168)**

| **Reference** | **Country** | **Age/sex** | **Duration of illness** | **Presenting complaints** | **Details of vision loss** | **Neuroimaging** | **Location of the cyst** | **Anatomical location of vision loss** | **Tissue biopsy** | **Medical treatment** | **Whether vision loss after albendazole** | **Surgery** | **Outcome** |
| --- | --- | --- | --- | --- | --- | --- | --- | --- | --- | --- | --- | --- | --- |
| Konar et al 2024 | India | 34/F | 2 months | NA | Complete vision loss on left side  Right hemianopia | Multiple cystic lesions in the suprasellar cistern | Subarachnoid NCC | Optic chiasma | Cysticercal larva with scolex | Albendazole and corticosteroids | NA | Excision of the cysts | Vision returned to normal. |
| Thangamathesvaran et al 2023 | India | 7/F | NA | NA | Left vision loss  Acuity= 20/25 | NA | Intraretinal  Ocular cysticercosis | Retina | NA | Albendazole and corticosteroids | NA | NA | NA |
| Rehman et al 2023 | USA  Immigrant | 44/F | 5 years | Headache  Memory loss | Progressive vision loss bilateral | A partially  calcified suprasellar/parasailer mass | Subarachnoid NCC | Optic chiasma | Cysticercal larva with scolex | Albendazole and corticosteroids | NA | Excision of the mass lesion | Vision progressively improved |
| Rama et al 2023 | India | 45/F | 15 days | Seizures | Right vision loss 6/24 | NA | Intravitreal cystic lesion with scolex  Retinal detachment | Retina | NA | Albendazole and corticosteroids | NA | Excision of the cysts | Vision returned to normal. |
| Mansour and Tahir 2023 | Egypt | 36/M | 2 weeks | Headache and vomiting  Papilledema | Bilateral vision loss  Right 6/12  Left 6/18 | Intraventricular cyst with scolex | Intraventricular NCC | Ventricles | Cysticercal larva with scolex | Albendazole and corticosteroids | NA | Excision of the cysts | Vision returned to normal. |
| Grover et al 2023 | India | 20/M | 1 month | NA | Vision loss right | Normal | Intravitreal cystic lesion with scolex  Retinal detachment | Retina | Cysticercal larva with scolex | Albendazole and corticosteroids | NA | Excision of the cysts | Vision returned to normal |
| Gasca-Sánchez et al 2023 | Colombia | 4/F | NA | NA | Vision loss right | Normal | Intravitreal cystic lesion with scolex with hemorrhage and retinal detachment  A a cystic lesion on the optic disc | Retina and optic disk, optic nerve | NA | NA | NA | Excision of the cysts | No significant improvement |
| Fan et al 2023 | USA | 40/M | 6 months | Headache  Elevated cells and protein in CSF and presence of eosinophilia | Initially 20/20 but had papilledema  One month later 20/40 on right side | NA | Intramedullary spinal cord NCC | Spinal NCC  Eosinophilic meningitis | Biopsy of spinal lesion= Cysticercal larva with scolex | Albendazole and corticosteroids | NA | Excision of the spinal cysts | Vision returned to normal |
|  |  | 40/F | 4 years | Headache  Diplopia  Elevated cells in CSF and presence of eosinophilia | Transient vision loss  Initially 20/20 but had papilledema | Racemose NCC in the in the chiasmatic cistern | Subarachnoid NCC | Optic chiasma | Metagenomic next-generation sequencing= *Teania solium* | Albendazole, praziquantel and corticosteroids | NA | NA | Condition was stable. |
| Das et al 2023 | India | 6/M | 2 months | Pain and redness of eyes | Vision loss bilateral limited to light perception on right side  Left= 20/30 | A calcified lesion of the brain | Ocular NCC  Retinal cyst with scolex and retinal detachment | Retina | Cysticercal larva with scolex | Albendazole and corticosteroids | NA | Excision of the eye ball | Vision returned to normal. |
| Burgos-Sosa et al 2023 | Mexico | 35/M | 1 month | Headache  Mental changes | Vision loss | A cystic lesion in sellar and suprasellar region | Subarachnoid NCC | Optic chiasma | Cysticercal larva with scolex | Albendazole and corticosteroids | NA | Excision of the cysts | Vision improved |
|  |  | 52/M | 4 months | NA | Vision loss | A cystic lesion in sellar and suprasellar region | Subarachnoid NCC | Optic chiasma | Cysticercal larva with scolex | Albendazole and corticosteroids | NA | Excision of the cysts | Vision improved |
|  |  | 46/F | 5 months | Headache  Gait abnormality  Seizures | Vision loss | Cystic lesions in frontal and temporal region compressing over suprasellar region | Subarachnoid NCC | Optic chiasma | Cysticercal larva with scolex | Albendazole and corticosteroids | NA | Excision of the cysts | Vision improved |
| Bansal et al 2023 | India | 18/M | 1 week | Headache | Vision loss  Acuity =20/30 | Disseminated innumerable cysts | Disseminated NCC  Subretinal cyst with a scolex | Retina | NA | NA | NA | Excision of the retinal cyst | NA |
| Koonwar et al 2022 | India | 1/M | 1 week | Fever  Seizure  Altered sensorium | Cortical blindness  Visual abnormality was noticed after he regained consciousness | Disseminated innumerable cysts | Disseminated NCC | Visual cortex | NA | Albendazole and corticosteroids | NA | NA | Vision not improved |
| Opara 2022 | USA | 14/M | 3 days | Fever  Seizure  Altered sensorium  Hemiparesis  Elevated cells and protein in CSF | Vision loss bilateral  Cortical blindness | Multiple cystic lesions with scolex  Cerebral infarcts | Parenchymal NCC | Visual cortex | NA | Albendazole and corticosteroids | NA | NA | Vision partially improved  Brain imaging became normal. |
| Jethva et al 2022 | India | 55/F | 1 year | Subcutaneous nodules  Difficulty in walking | Bilateral vision loss  Right no perception of light  Left 6/12  RAPD right | Numerous cystic lesions in the brain | Left eye a subconjunctival cyst  Right Intravetreous cyst with scolex | Retina  Disseminated NCC | NA | Albendazole and corticosteroids | NA | NA | NA |
| Li et al 2022 | USA | 11/M | Sudden | NA | Vision loss right  20/200  Patient had chronic posterior uveitis from 3 years.  Multifocal subretinal fluid | NA | Ocular NCC  Multifocal subretinal fluid and retinal detachment  Intravetreous cyst | Retina | Cysticercal larva with scolex | Albendazole and corticosteroids | NA | Excision of the cysts | Vision improved |
| Koirala et al 2022 | Nepal | 27/M | Sudden | Diplopia  Periorbital pain | Vision loss left 6/60  RAPD left eye  6^th^ nerve palsy left | A cystic lesion in left rectus muscle in orbital apex compressing over optic nerve | Orbital apex syndrome | Optic nerve | NA | Albendazole and corticosteroids | NA | NA | Vision improved |
| Shrestha et al 2021 | Nepal | 42/F | 1 month | NA | Vision loss right  5/60  No vision in left eye over 10 years  RAPD left eye | Calcified lesions in the brain | Subretinal cyst | Retina | Cysticercal larva with scolex | Albendazole and corticosteroids | NA | Excision of the cyst | Vision improved |
| Shakya et al 2021 | India | 28/M | NA | Headache | Vision loss left | A cystic lesion in suprasellar region | Subarachnoid NCC | Optic chiasma | Cysticercal larva with scolex | NA | NA | Excision of the cyst | Vision improved |
| Samanta et al 2021 | India | 10/F | 3 weeks | Headache | Vision loss right | Multiple cystic lesions | Subretinal cysticercosis | Retina | NA | Oral corticosteroids | NA | NA | Vision not improved |
| Niang et al 2021 | Senegal | 12/M | 6 months | Seizures  Headache | Vision loss | Innumerable small cysts in brain | Cysticercosis encephalitis | Cerebral cortex | NA | Corticosteroids | NA | NA | NA |
| Kumar and Padhy 2021 | India | 22/M | 1 month | Headache | Vision loss right 20/400 | Innumerable small cysts in brain | Cyst in vitreous fluid  Sub macular cysticercus | Retina | NA | NA | NA | Excision of the cyst | Vision improved |
| Gala et al 2021 | India | 10/M | 1 week | Fever  Headache and vomiting  Seizure  Altered sensorium  Elevated cells and protein in CSF | Progressive loss of vision over 2 months following albendazole treatment | Innumerable small cysts in brain  Hydrocephalus | Cysticercosis encephalitis  Optic atrophy | Cerebral cortex | NA | Corticosteroids | NA | NA | NA |
| Bhatia et al 2021 | India | 60/M | 2 years | Seizures  Mental decline  Skin nodule | Vision loss right | Innumerable small cysts with scolex in brain and other body parts | Disseminated NCC | Ocular NCC part of Disseminated NCC | Biopsy of skin nodule= Cysticercal larva with scolex | Corticosteroids | NA | NA | Improved |
| Snyder et al 2020 | USA | 39/M | 2 months | Headache | Vision loss bilateral right 1/200E | A cystic lesion with partial calcification in suprasellar region compressing over optic chiasma | Subarachnoid NCC | Optic chiasma | Cysticercal larva with scolex | NA | NA | Excision of the cyst | Improved |
| Silva et al 2020 | Brazil | 46/F | 2 weeks | Recurrent syncope and falls  Bruns syndrome  Seizures | Progressive vision loss | Cysts in forth ventricle with hydrocephalus | Ventricular NCC | Optic chiasma | NA | Albendazole and corticosteroids | NA | Excision of the cyst | Died |
| Shashni et al 2020 | India | 34/M | Sudden | NA | Vision loss left 1/60 | NA | Subretinal  Cyst with scolex | Retina | NA | NA | NA | Excision of the cyst | NA |
| Samanta et al 2020 | India | 55/M | NA | NA | Vision loss left 3/60 | NA | Subretinal Intravetreous  cyst with scolex  Retinal detachment | Retina | NA | NA | NA | Excision of the cyst | Not improved |
|  |  | 35/F | NA | NA | Vision loss left 6/60 | NA | Subretinal Intravetreous  cyst with scolex  Retinal detachment | Retina | NA | NA | NA | NA | NA |
| Reddy et al 2020 | India | 31/F | Sudden | NA | Vision loss right 20/600 | NA | Subretinal Intravetreous  cyst with scolex  Retinal detachment | Retina | NA | Anthelminthic drug and corticosteroids | NA | Excision of the cyst | NA |
| Patidar et al 2020 | India | 23/M | 20 days | Ptosis right | Vision loss right  Primary  optic atrophy | A cystic lesion in superior rectus muscle within optic canal | Orbital apex syndrome | Optic nerve | Cysticercal larva with scolex | Albendazole and corticosteroids | NA | Excision of the cyst | NA |
| Neto et al 2020 | Brazil | 39/M | 10 days | NA  CSF cells increased | Vision loss right 20/400  Chorioretinitis | An enhancing lesion in the optic nerve head | Retrobulbar lesion | Optic nerve | NA | Corticosteroids | NA | NA | Vision improved |
| Lee et al 2020 | Korea | 52/M | 2 weeks | Headache | Vision loss right  Right homonymous hemianopia | An enhancing lesion in the left occipital lobe | Colloidal lesion | Occipital cortex | Cysticercal larva with scolex | Albendazole and corticosteroids  Patient did not respond to medical treatment. | NA | Excision of the cyst | Vision improved |
| Kumar et al 2020 | India | 53/M | 2 weeks | NA | Vision loss left 6/60 | Normal | A large sub macular  cysticercosis cyst with scolex | Retina | NA | Corticosteroids | NA | Excision of the cyst | Vision improved |
| García Franco et al 2020 | Mexico | 25/F | 3 months | NA | Vision loss left 20/100 | Normal | A large sub macular  cysticercosis cyst with scolex | Retina | NA | Albendazole and corticosteroids | NA | NA | Vision improved |
| Dhiman et al 2020 | India | 28/F | 1 week | Headache  Diplopia | Vision loss right 6/12  RAPD | A ring enhancing lesion with scolex in right medial rectus muscle | Orbital apex syndrome | Optic nerve | NA | Albendazole and corticosteroids | NA | NA | Vision improved |
| Cuellar-Hernandez et al 2020 | Mexico | 45/F | 6 moths | Headache | Vision loss left 20/80  Bitemporal hemianopia | A cystic lesion in sellar and suprasellar region | Subarachnoid NCC | Optic chiasma | Cysticercal larva with scolex | NA | NA | Excision of the cyst | Vision improved |
| Zou et al 2019 | China | 18/F | 1 day | Fever  Headache | Vision loss left 20/40  A cyst in the left vitreous cavity  Retinal vasculitis with optic disc edema | Innumerable lesion throughout the body including brain | Disseminated NCC | Retina and optic nerve | Cysticercal larva with scolex | Albendazole and corticosteroids | NA | Excision of the cyst | Vision improved  Number of lesion reduced throughout the body |
|  |  | 24/M | NA | Headache | Vision loss left 20/40 | Innumerable lesion throughout the body including brain | Disseminated NCC | Extraocular muscle affecting optic nerve | Cysticercal larva with scolex | Albendazole and corticosteroids | NA | Excision of the cyst | Vision improved  Number of lesion reduced throughout the body |
|  |  | 42/M | NA | Seizure | Vision loss right | Innumerable lesion throughout the body including brain | Disseminated NCC | Superior rectus muscle affecting optic nerve | Cysticercal larva with scolex | Albendazole and corticosteroids | NA | Excision of the cyst | Vision improved  Number of lesion reduced throughout the body |
| Wadhwani et al 2019 | India | 24/M | 1 week | NA | Vision loss bilateral no light perception | Innumerable lesion throughout the brain  Lesions in both optic nerve sheath | Disseminated NCC | Optic nerve | NA | Corticosteroids | NA | NA | Not improved |
| Shrestha and Shrestha 2019 | Nepal | 22/M | 1 month | Headache  Skin nodules | Vision loss bilateral 6/12 in both the eyes  Papilledema | Innumerable lesion throughout the brain | Disseminated NCC | Optic nerve | NA | Albendazole and corticosteroids | NA | NA | NA |
| Musara et al 2019 | Zimbabwe | 35/M | 2 weeks | Headache | Vision loss right | A cystic lesion in sellar and suprasellar region | Subarachnoid NCC | Optic chiasma | Cysticercal larva with scolex | Albendazole and praziquantel and corticosteroids | NA | Excision of the cyst | Vision improved |
| Kumar et al 2019 | India | 17/F | 3 months | NA |  | Normal | Subretinal Intravetreous  cyst with scolex  with retinal fibrosis  Ocular NCC | Retina | NA | NA | NA | Excision of the cyst | Vision not improved |
| Chaugule et al 2019 | India | 22/M | Sudden | NA | Vision loss left  RAPD  Ophthalmoplegia | A cystic  lesion with scolex and enhancement within the optic nerve at the orbital apex | Orbital apex syndrome | Optic nerve | NA | Albendazole and corticosteroids | NA | NA | Vision improved and lesion decreased |
| Swamy et al 2018 | India | 26/F | Sudden | NA | Vision loss left no light perception | Normal | A total retinal detachment with a  subretinal cyst with  scolex | Retina | NA | Albendazole and corticosteroids | NA | NA | Vision not improved but lesion decreased |
| Mukhija et al 2018 | India | 26/M | 1 month | NA | Vision loss left 6/60 | NA | A cystic lesion in the left orbit extending from the orbital apex  Orbital apex syndrome | Optic nerve | NA | Corticosteroids | NA | NA | Improved |
| Matalia et al 2018 | India | 20/F | 3 days | NA | Vision loss left 6/18  RAPD | NA | A cystic lesion with scolex in lateral rectus muscle and encroaching orbital apex | Optic nerve | NA | Albendazole and corticosteroids | NA | NA | Vision improved |
| Goel 2018 | India | 33/F | 1 month | Headache | Vision loss right hand movement perception  Disk edema  RAPD | NA | A cystic lesion  with enhancement in the optic nerve | Optic nerve | NA | Albendazole and corticosteroids | NA | NA | Vision improved |
| Obedulla et al 2017 | India | 6/M | 10 days | NA | Vision loss left 20/60  Conjunctival swelling  Ophthalmoplegia | NA | Spontaneous extrusion of the subconjunctival  Cyst  Ocular NCC | Anterior chamber | NA | Albendazole and corticosteroids | NA | NA | NA |
| Majumdar and Pal 2017 | India | 20/M | NA | NA | Vision loss left 20/80 | NA | A large intravitreal cyst  Retinal detachment, and extensive proliferative  vitreoretinopathy | Retina | NA | NA | NA | NA | NA |
| Karthikeya et al 2017 | India | 34/M | 6 months | NA | Vision loss right  3/60 | Multiple ring-enhancing lesions of the brain | Intravitreal cysticercosis with scolex | Vitreous fluid entering through a break in the retina | NA | NA | NA | Excision of the cyst | Vision improved |
| Gaur et al 2017 | India | 18/M | 2 months | Headache | Vision loss bilateral perception light only | Miliary lesions of NCC  Some lesions with scolex and perilesional edema | Parenchymal NCC  Lesions near optic chiasma  Occipital lobe lesions revealed surrounding edema | Primary optic atrophy and  cortical visual loss  Optic nerve, optic chiasma and visual cortex | NA | Anthelminthic drug and corticosteroids | NA | NA | Not improved |
| Singh et al 2016 | India | 18/F | 15 days | NA | Vision loss right  20/80  6/36 in both eyes  Conjunctival inflammation | NA | Two cysts with scolex in the anterior  chamber on right side | Anterior chamber | Cysticercal larva with scolex | Albendazole and corticosteroids | NA | Excision of the cyst | Vision improved |
| Nie et al 2016 | China | 6/F | NA | NA | Vision loss left  Strabismus | A large cyst in eft cerebral  hemisphere with no perilesional edema and contrast enhancement | Cerebral cortex | Raise ICT  Optic nerve | Cysticercal larva with scolex | Anthelminthic drug and corticosteroids | NA | Excision of the cyst | NA |
| Krupa et al 2016 | USA | 39/M | 1 month | Headache  CSF = increasing cells and protein | Vision loss  Bilateral papilledema | Innumerable confluent cystic lesions in the the suprasellar, interpeduncular, and prepontine cisterns cisterns and sylvian fissures, lateral ventricles  a small subarachnoid cystic lesion at the level of the conus  Hydrocephalus | Racemose subarachnoid NCC | Optic nerve and optic chiasma | NA | Albendazole and corticosteroids | NA | VP shunt | Improved  Complete resolution of the lesions |
| Khalid et al 2016 | India | 17/M | 20 days | Headache | Vision loss  RAPD left eye  Bilateral papilledema | Fourth ventricular cyst that later migrated to third ventricle  Hydrocephalus | Intraventricular NCC | Raised ICT  Optic nerve | NA | Albendazole and corticosteroids | NA | VP shunt | Improved |
| Jain et al 2016 | India | 23/F | 1 month | Orbital pain | Vision loss left no light perception | Cyst in retroorbital left optic nerve region  Cyst with scolex in left medial rectus | Ocular NCC | Optic nerve | NA | Albendazole and corticosteroids | NA | NA | Improved |
| Wang and Huang 2015 | USA | 45/M | 4 years | Headache | Vision loss  Bilateral optic atrophy | Obstructive hydrocephalus  A mass lesion compressing optic  chiasma, Sella, and midbrain abutting corticomedullary junction | Racemose subarachnoid NCC | Optic chiasma | NA | Albendazole and corticosteroids | NA | NA | Headache improved but vision remained unaltered |
| Vaitheeswaran et al 2015 | India | 32/M | 3 days | Headache | Vision loss right 3/60  RAPD | A cystic lesion with scolex in right optic nerve  Innumerable brain lesions | Parenchymal NCC | Optic nerve | NA | Albendazole and corticosteroids | NA | Excision of the cyst | Vision improved |
| Singh and Singh  2015 | India | 24/F | 1 week | NA | Vision loss right 5/300  RAPD | Normal | Subretinal lesion at macula retinal hemorrhage and with retinal detachment | Retina | NA | Albendazole and corticosteroids | NA | NA | Vision improved |
| Sharma et al 2015 | India | 17/M | 2 weeks | NA | Episodic vision loss left | NA | Intraretinal cyst with retinal detachment | Retina | NA | Albendazole and corticosteroids | NA | NA | Enucleation of the eye |
| Sharma et al 2015 | India | 17/M | 3 months | Headache  Diplopia  Cerebellar signs | Vision loss left 6/24 | Multiple ring enhancing lesions of the brain  Disseminated NCC | A ring lesion in left lateral rectus | Raised ICT  Optic nerve | NA | Anthelminthic drug and corticosteroids | NA | NA | Improved |
| Nehasudhakar et al 2015 | India | 10/F | 5 days | NA | Vision loss right no perception of light | Multiple calcified lesions of the brain | Intravetreous cyst with scolex  Ocular NCC | Retina | NA | Anthelminthic drug and corticosteroids | NA | NA | NA |
| Narra et al 2015 | India | 21/M | 1 month | Headache | Vision loss left  Left optic disk edema | NA  A small cyst in left temporalis muscle | Cyst in left optic nerve  Ocular NCC | Optic nerve | NA | Anthelminthic drug and corticosteroids | NA | NA | Marked improvement in vision |
| Mishra et al 2015 | India | 29/F | 9 months | NA | Vision loss left 1/60 | NA | Intravetreous cyst with scolex  Retinal hemorrhage and detachment | Retina | NA | Anthelminthic drug and corticosteroids | NA | NA | Marked improvement in vision |
| Jain et al 2015 | India | 14/F | 5 months | Seizures | Vision loss bilaterally | Innumerable cystic lesion in whole of the brain  Disseminated NCC | Subretinal cysts  Retinal hemorrhage and retinal detachment | Retina | Cysticercal larva with scolex | Corticosteroids | NA | Excision of the cyst was possible from right eye, in left cyst get ruptured | Vision not improved |
| Huang and Sridhar 2015 | USA | 30/F | 3 months | Tinnitus | Bilateral  blurry vision with transient visual obscurations  Papilledema bilateral | A large 4^th^ ventricular cyst  Innumerable cystic lesion in whole of the brain  Hydrocephalus | Ventricular NCC | Raised ICT  Optic nerve | Cysticercal larva with scolex | Albendazole and corticosteroids | NA | Ventriculoperitoneal  Shunt  Surgical excision of ventricular cyst | Marked improvement in vision |
| Chavala et al 2015 | USA | 25/M | 6 months | NA | Vision loss left perception of light | NA | Intraretinal cyst | Retina | NA | Albendazole | NA | NA | Improved |
| Chaudhary et al 2015 | India | 8/M | 6 months | Headache | Vision loss bilateral limited to finger counting  Optic atrophy | Innumerable enhancing lesions in whole of the brain  Hydrocephalus | Parenchymal NCC | Optic nerve  Raised ICT | NA | Albendazole and corticosteroids | NA | Ventriculoperitoneal  Shunt | Marked improvement in vision |
| Wani et al 2014 | Kuwait | 24/M | 20 days | NA | Vision loss Left eye vision loss at 1/3 m | Innumerable cystic lesions in the brain | A subretinal translucent cyst with scolex | Retina | Cysticercal larva with scolex | Albendazole and corticosteroids | NA | Excision of cysts | Vision improved |
| Tran et al 2014 | USA | 29/F | 4 months | Headache  Somnolence | Bilateral vision loss | Cysts in the fourth ventricle | Intraventricular | Ventricle | NA | Albendazole and corticosteroids | NA | Excision of cysts | Vision improved |
| Takkar et al 2014 | India | 6/F | NA | NA | Vision loss left eye | NA | Ocular | Anterior Chamber of Eye | NA | Topical corticosteroids | NA | Excision of cyst | Vision improved |
| Sune et al 2014 | India | 40/F | 2 months | Mental changes  Headache | Bilateral vision loss  20/125 | Innumerable cystic lesions with scolex in the brain  colloid vesicular stage NCC | Cysticercosis encephalitis | Raised ICT  Optic nerve | NA | Albendazole, praziquantel and corticosteroids | NA | NA | Vision improved to 20/40 |
| Salcedo-Villanueva et al 2014 | Mexico | 45/F | 5 years | Seizures  Headache and vomiting | Vision loss bilateral  20/40  Bitemporal heteronymous hemianopia. | Basal arachnoiditis and hydrocephalus | intraventricular vesicular lesion in the 3rd ventricle | Ventricle | NA | Albendazole and corticosteroids | NA | VP shunt | NA |
| Mohan et al 2014 | India | 53/M | NA | NA | Vision loss | NA | Ocular cysticercosis -macular subretinal cystoid abnormality | Retina | NA | NA | NA | Excision of cyst | Vision improved to 20/400 |
| Matalia et al 2014 | India | 22/M | 4 months | Headache | Vision loss bilateral  Right no perception of light  Left 6/12  Papilledema | Multiple lobulated cysts without scolex a racemose NCC in basal cisterns  Hydrocephalus | Subarachnoid NCC | Optic Chiasma | Cysticercal larva with scolex | Albendazole and corticosteroids | NA | Excision of cyst | Vision improved in left eye  No change in right eye |
| Kasundra et al 2014 | India | 41/M | Sudden | NA | Vision loss Right no perception of light | Innumerable cystic lesions with scolex in the brain | Subretinal choroidal lesions | Retina  Disseminated NCC | NA | Corticosteroids | NA | NA | No Improvement in vision |
| Das et al 2014 | India | 22/M | 7 days | NA | Vision loss right 20/60  Proptosis  Orbital cellulitis  Ophthalmoplegia | A cystic lesion with scolex in left occipital lobe | A subretinal cyst | Retina | NA | Albendazole and corticosteroids | NA | NA | Improvement in vision |
| Ko et al 2013 | Korea | 42/F | 4 months | NA | Vision loss right  finger counting | NA | Intravitreal cyst | Retina | NA | NA | NA | Excision of cyst | Vision improved to 0.5 |
| Kori et al 2013 | India | 31/F | 2 weeks | Headache | Vision loss right  finger counting | A cystic lesion with scolex in left frontal region | Subretinal cyst | Retina | NA | Corticosteroids | NA | Laser Photocoagulation | NA |
| Babalola et al 2013 | Nigeria | 32/M | 2 months | Headache | Vision loss right | NA | A subretinal cyst  The choroidal detachment | Retina | NA | Albendazole | Developed retinal hemorrhage | NA | NA |
| Aulakh 2013 | India | 8/F | 10 days | Headache  Seizures  Altered consciousness | Vision loss right | Multiple enhancing lesions with severe perilesional edema | Parenchymal NCC | Cortical blindness | NA | Albendazole and corticosteroids | NA | NA | Vision not improved  She became fully conscious |
| Venna et al 2012 | USA | 42/F | 2 weeks | Headache  Diplopia | Vision loss right 20/50  Bilateral papilledema | 4^th^ Ventricle    Spinal meninges | Raised ICT | Intraventricular NCC | Cysticercal larva with scolex | Albendazole, praziquantel and corticosteroids  6 months treatment | NA | Excision of cyst | Vision improved |
| Swastika et al 2012 | Indonesia | 9/F | 2 weeks | NA | Vision loss left 6/10 | NA | Ocular cysticercosis | anterior chamber of the left eye | Cysticercal larva with scolex | Topical corticosteroids | NA | Excision of cyst | Vision improved |
| Sinha S et al 2012 | India | 12/M | NA | NA | Vision loss 4/60 | NA | Subretinal NCC  Ocular cysticercosis | Retina | Cysticercal larva with scolex | NA | NA | Vitrectomy with cyst aspiration | NA |
| Raval et al 2012 | India | 31/M | 6 months | NA | Vision loss right  light perception in right eye | Ring- enhancing lesion in left cerebellar hemisphere with perilesional oedema | Subretinal cyst | Retina | NA | Albendazole and corticosteroids | NA | NA | NA |
| Prasad et al 2012 | India | 7/M | 1 month | Headache and vomiting | Vision loss left  Proptosis | Multiple cystic lesions in whole brain | An elevated cystic mass over macula | Retina  Disseminated NCC | NA | Albendazole and corticosteroids | Cysticidal  syndrome | No | Improved |
| Parashari et al 2012 | India | 10/M | NA | NA | Vision loss left  6/9 | A cyst in right parietal region | Intravetreous cyst | Retina | NA | NA | No | Excision of cyst | NA |
| Mulla et al 2012 | India | 18/M | 1 year | NA | Bilateral vision loss  Endogenous endophthalmitis in the left eye | NA | Intravetreous cyst right | Retina | Cysticercal larva with scolex | Topical corticosteroids | NA | Excision of cyst | NA |
| Hussain et al 2012 | United Arab Emirates | 5/F | Sudden | NA | Episodic vision loss both side | An enhancing lesion in left occipital lobe | Cortex | Cortical vision loss as an epileptic manifestation | NA | Albendazole  And  Antiepileptic drug | NA | NA | Improved, no recurrence |
| Agarwal et al 2012 | India | 14/M | 1 month | NA | Vision loss right 1/60 | Multiple cystic lesions of the brain | Intravitreal cyst  Retinal detachment  A subconjunctival cysticercus cyst in the left eye | Retina | Cysticercal larva with scolex | Albendazole and corticosteroids | An anaphylactic reaction | Excision of cyst | Vision not improved |
| Sundar et al 2011 | India | 32/M | 2 weeks | Seizures | Vision loss Left light perception | Multiple cystic lesions of brain | Subretinal cyst  Retinal detachment | Retina | NA | Albendazole and corticosteroids | NA | Excision of cyst | NA |
| Ratra et al 2010 | India | 26/M | 15 days | NA | Vision loss right 20/200no light perception | NA | Intravitreal cysticercosis  Total retinal detachment | Retina | Cysticercal larva with scolex | albendazole | NA | Excision of cyst | Vision improved |
|  |  | 18/M | NA | NA | Vision loss left 20/40 | NA | Intravitreal cysticercosis  Retinal detachment | Retina | Cysticercal larva with scolex | NA | NA | Excision of cyst | Vision improved |
| Prasad et al 2010 | India | 6/M | 5 days | Fever  Headache  Seizures | Vision loss bilateral perception of light only | Multiple calcified lesions of the brain | Cysticercosis encephalitis | Cortical vision loss | NA | Albendazole and corticosteroids | NA | NA | Improved |
| Dass et al 2010 | India | 8/NA | 3 years | Seizure | Vision loss perception of light  Bilateral optic nerve atrophy | Multiple cystic lesions of the brain | 4^th^ ventricular cyst wit hydrocephalus | Optic nerve | NA | Corticosteroids | NA | Ventriculoperitoneal shunt | Improved |
| Cheong et al 2010 | Korea | 71/F | 4 years | NA | Vision loss bilateral | A cystic lesion in Pituitary Stalk | NA | Optic nerve | Cysticercal larva with scolex | NA | NA | Excision of cyst | Vision not improved |
| Yadav et al 2009 | Nepal | 22/M | 2 months | NA | Vision loss left 6/24 | NA | Intravetreous cyst | Retina | NA | Albendazole and corticosteroids | NA | Excision of cyst | Improved |
|  |  | 35/F | NA | NA | Vision loss right hand movements | NA | Intravetreous cyst | Retina | NA | Albendazole and corticosteroids | NA | Excision of cyst | Improved |
|  |  | 25/M | 1 month | NA | Vision loss left finger counting | Multiple cystic lesions of the brain | Intravetreous cyst | Retina | NA | Albendazole and corticosteroids | NA | Excision of cyst | Improved |
| Taksande et al 2009 | India | 40/M | 7 days | NA | Vision loss left | Cysts in left Medial rectus and superior rectus muscles with compression of left optic nerve | Orbital apex syndrome | Optic nerve | NA | Albendazole and corticosteroids | NA | NA | Improved |
| Patnaik et al 2009 | Nepal | 35/M | 3 months | Headache    Altered behaviour | loss of vision bilateral | Numerous ring enhancing lesions with scolex bilateral in brain | Cysticercotic encephalitis  Disseminated NCC | Raised ICT  Optic nerve | Cysticercal larva with scolex | Albendazole and corticosteroids | NA | NA | Improved |
| Miller et al 2009 | USA | 24/M | 4 months | Headache | Loss of vision left 20/80 | Multiple grape like lesions in subarachnoid space  Multiple parenchymal cysts | Subarachnoid NCC | Optic chiasma | NA | Albendazole, corticosteroids | NA | NA | Improved |
| Knight et al 2009 | UK | 32/F | 5 days | Headache | Loss of vision bilateral  Right 6/36  Left 6/12 | 4^th^ ventricular cyst  Hydrocephalus | Ventricular NCC | Raised ICT  Optic nerve | Cysticercal larva with scolex  Multiple extra parenchymal grape-like cysts | VP shunt  Praziquantel and albendazole | NA | Excision of cyst | Improved |
| Venkatesh et al 2008 | India | 31/M | Sudden | NA | Recurrent vision loss right 20/40  Disk edema  RAPD  Next episode 3 years later | A cystic lesion within optic nerve | Ocular NCC | Optic nerve | NA | Albendazole and corticosteroids | NA | NA | Marked improvement in vision  Complete resolution of the lesion |
| Kai et al 2008 | India | 16/F | 7  months | NA | Vision loss left 1/60 | Multiple cystic lesions in the brain  Disseminated NCC | A live  cyst in the anterior chamber | Anterior chamber | Cysticercal larva with scolex | Albendazole and corticosteroids | NA | Excision of the cyst | Vision improved |
| Aghamohammadi et al 2008 | USA | 14/F | 2 months | NA | Vision loss left perception of light only  RAPD | Normal | Multiple cysts in the retina with retinal detachment | Retina | Cysticercal larva with scolex  *Taenia crassiceps* larvae | NA | NA | Excision of the cyst | NA |
| Shariq and Adhikari 2007 | Nepal | 5/F | 2 months | NA | Vision loss right 20/120 | Normal | A cyst in the anterior chamber of right side | Anterior chamber | Cysticercal larva with scolex | Corticosteroids | NA | Excision of the cyst | Vision improved |
| Mahendradas et al 2007 | India | 10/M | 15 days | Ocular pain and redness in left eye  Severe headache | Vision loss left 20/40 | Normal | A cyst in the anterior chamber of left side | Anterior chamber | Cysticercal larva with scolex | NA | NA | Excision of the cyst | Vision improved |
| Karande and Kumbhare 2007 | India | 5/M | NA | Seizures  1 year later  Ocular inflammation | Vision loss left no perception of light | Multiple cystic lesions of the brain | A cystic lesion  along the posterior retina  Retinal detachment | Retina  Disseminated NCC | NA | Albendazole and corticosteroids | Inflammatory  phthisis and lost all vision in  his left eye | NA | No improvement  the child developed inflammatory  phthisis |
| Goyal et al 2007 | India | 10/M | 2 months | NA | Vision loss right no perception of light  Proptosis | NA | A cystic lesion in the right inferior rectus muscle, extending  up to the orbital apex | Orbital apex syndrome  Optic nerve | NA | Albendazole and corticosteroids | NA | NA | Vision improved and proptosis resolved |
| Cortez et al 2007 | Colombia | 6/F | NA | NA | Vision loss left limited to perception of hand movement | NA | A cyst n the anterior chamber and scolex was noted | Anterior chamber | Cysticercal larva with scolex | NA | NA | Excision of the cyst | Vision improved |
| Sudan et al 2005 | India | 32/F | 4 weeks | NA | Vision loss left 6/60  RAPD | Normal | A cystic lesion with scolex in the optic nerve  head | Optic nerve | NA | Albendazole and corticosteroids | NA | NA | Vision improved and lesion resolved |
| Pushker et al 2005 | India | 50/F | 1 month | Headache and vomiting  Subcutaneous nodules | Vision loss bilateral 20/40 right and 20/50 left. | Innumerable small cystic lesions | an orbital cyst with  scolex in the right orbit | Disseminated NCC  Raised ICT leading to optic nerve damage | NA | Albendazole and corticosteroids | NA | NA | Vision improved |
| Chadha et al 2005 | India | 21/M | 2 months | Seizures | Vision loss right  No perception of light  RAPD | NA | An intravitreal cyst with exudative retinal detachment  Cyst present in ocular muscles as well | Retina | NA | Albendazole and corticosteroids | NA | NA | Vision improved |
| Besada et al 2005 | USA | 41/F | NA | Headache | Vision loss bilateral right 20/25  Left 20/30  Bilateral disk edema | A large cystic lesion in left temporal lobe compressing over optic chiasma | A retinal lesion and brain lesion | Retina and optic chiasma | NA | Albendazole | NA | NA | Vision improved only on left side |
| Lim and Chee 2004 | Singapore | 51/M | 6 months | NA | Vision loss right | Multiple enhancing lesions of the brain | A subretinal cystic lesion | Retina | NA | Albendazole and corticosteroids | NA | NA | Initially vision deteriorated as patient developed uveitis.  Vision improved |
| Bowie et al 2004 | USA | 38/M | 9 months | Headache  Seizure | Vision loss bilateral  right 20/70  Left 20/100  Serous  retinal detachments | Multilocular cystic  lesions in the lateral ventricle  Hydrocephalus | Intraventricular NCC | Retina and optic nerve | NA | Albendazole and corticosteroids | NA | NA | Vision improved |
| Hiralal et al 2003 | India | 17/F | 2 months | NA | Vision loss right  Proptosis | Normal | A cystic lesion with scolex in right optic nerve | Optic nerve | NA | Albendazole and corticosteroids | NA | NA | Vision improved |
| Adegbehingbe et al 2003 | Nigeria | 43/F | 18 months | NA | Vision loss left perception of light | NA | Intravetreous cyst with scolex  Ocular NCC | Retina | NA | Praziquantel | NA | NA | NA |
| Verma et al 2002 | India | 30/M | 10 days | NA | Vision loss left  Disk edema  RAPD | NA | Optic nerve cyst | Optic nerve | NA | Corticosteroids | NA | NA | Vision improved |
| Das et al 2002 | India | 74/M | 3 months | NA | Vision loss bilateral  Right 6/60  Left 6/12 | NA | A cystic lesion in anterior chamber  Lesion with scolex was protruding in posterior chamber as well | Anterior chamber | Cysticercal larva with scolex | Albendazole and corticosteroids | NA | Excision of the cyst | Vision improved |
| Chung et al 2002 | USA | 56/F | 1 month | NA | Vision loss left  20/200  RAPD  Vitreous hemorrhage | An enhancing lesion in occipital lobe | A cystic lesion with scolex in subretinal region  Retinal detachment | Retina | NA | Anthelminthic drug and corticosteroids | NA | Excision of the cyst | NA |
| Bajaj and Pushker 2002 | India | 24/F | Sudden | NA | Vision loss right  Proptosis  These symptoms were relieved on treatment  Second episode 16 | A cystic lesion with scolex in right optic nerve | NA | Optic nerve | NA | Albendazole and corticosteroids | NA | NA | Vision improved  Complete resolution of the cyst |
| Sidhu et al 2001 | India | 25/F | 3 months | NA | Vision loss left | NA | A cystic lesion with scolex in vitreous cavity  Retinal detachment | Retina | NA | NA | NA | NA | NA |
| Sabti et al 2001 | Kuwait | 35/M | 4 days | NA | Vision loss right  Later left side | NA | A subretinal cystic lesion with scolex  Retinal detachment  Cysts were noted on both sides | Retina | NA | Corticosteroids | NA | NA | Not improved |
| Lombardo 2001 | USA | 21/F | 1 week | NA | Vision loss right  20/400 | NA | A cystic subretinal lesion  Retinal hemorrhage | Retina | NA | Albendazole and corticosteroids | NA | NA | Lost to follow up |
| Chandra et al 2000 | India | 50/F | 6 months | Headache and ocular pain | Vision loss left  Proptosis | NA | A cystic lesion in left optic nerve | Optic nerve | NA | Albendazole and corticosteroids | NA | NA | Not improved |
| Gurha et al 1999 | India | 15/F | 6 months | Headache | Vision loss left  Only finger counting  RAPD  Papillitis | NA | A cystic lesion in left optic nerve | Optic nerve | Cysticercal larva with scolex | NA | NA | Excision of the cyst | Initially developed ptosis  Vision improved |
| Betharia et al 1999 | India | 15/M | 8 months | NA | Vision loss left  Only finger counting | NA | A cystic lesion in left optic nerve | Optic nerve | Cysticercal larva with scolex | Albendazole and corticosteroids | NA | Excision of the cyst | Vision improved |
| Tandon et al 1998 | India | 26/M | Sudden | NA | Diplopia  Vision loss left  20/80 5 days after albendazole | NA | A cystic lesion in left superior rectus muscle close to the orbital apex | Optic nerve | NA | Albendazole and corticosteroids | Optic neuritis | NA | Improved with continued corticosteroids |
|  |  | 32/M | 1 month | NA | Proptosis  Vision loss right  one week days after | NA | A cystic lesion in right orbital apex | Optic nerve | NA | Albendazole and corticosteroids | Optic neuritis | NA | Improved with continued corticosteroids |
| Gupta et al 1998 | India | 14/M | 2 months | NA | Vision loss left | NA | A cystic lesion in subretinal region | Retina | Cysticercal larva with scolex | NA | NA | Excision of the cyst | Vision not improved |
| George et al 1998 | India | 20/M | 2 months | NA | Vision loss left  6/18 | NA | A cystic Intravetreous lesion | Retina | NA | NA | NA | Excision of the cyst | Vision improved |
| Seo et al 1996 | Korea | 36/M | NA | NA | Vision loss right | NA | Intravitreal cysticercosis with scolex with retinal hemorrhages and retinal detachment | Retina | NA | Praziquantel and corticosteroids | NA | Excision of the cyst | Vision improved |
| Bousquet et al 1996 | France | 12/M | NA | NA | Vision loss left  4/10  Left disk edema | NA | A cystic lesion in left intraorbital optic nerve | Optic nerve | Cysticercal larva with scolex | Praziquantel and corticosteroids | NA | Excision of the cyst | Vision improved |
| Santoyo et al 1991 | Mexico | 43/F | 10 months | NA | Bilateral vision loss  Left identify objects at 30 cm  20/25 in right eye | Three large cysts in parasellar region | NA | Optic nerve and optic chiasma | NA | Albendazole and corticosteroids | NA | NA | Vision improved |
| Mason et al 1991 | Zimbabwe | 16/F | Sudden | NA | Vision loss left | A calcified lesion in occipital lobe | A cyst in anterior chamber | Anterior chamber | Cysticercal larva with scolex | Praziquantel and corticosteroids | NA | Excision of the cyst | Vision improved |
| Luger et al 1991 | The Netherlands | 44/M | 2 months | NA | Vision loss left  20/30 | NA | A subretinal cyst | Retina | Cysticercal larva with scolex | NA | NA | Excision of the cyst | Vision improved |
| Steinmetz et al 1989 | USA | 39/F | 2 weeks | NA | Vision loss left  20/40 | NA | A subretinal cyst with scolex  Foveal detachment | Retina | Cysticercal larva with scolex | NA | NA | Excision of the cyst | Vision improved |
| Pansey et al 1989 | India | 5/F | 6 months | Severe headache | Bilateral vision loss  Optic atrophy | Multiple enhancing lesions of the brain | NA | Raised ICT  Optic nerves | NA | Praziquantel and corticosteroids | NA | NA | Vision improved |
| Rafael and Gomez-Llata 1985 | France | 31/M | 7 months | NA | Bilateral vision loss finger counting only | Multiple cystic lesions in parasellar region of the brain | Subarachnoid NCC | Optic chiasma | NA | NA | NA | Excision of the cyst | Vision improved |
| Kruger-Leite et al 1985 | USA | 50/F | 3 months | NA | Vision loss left  Hand movement at 30 cm | NA | A subretinal Intravetreous cyst with scolex in macular region | Retina | NA | NA | NA | Excision of the cyst | Vision improved |
| Kestelyn and Taelman 1985 | Belgium | 45/M | 6 months | Subcutaneous nodules | Vision loss right light perception only | NA | A subretinal Intravetreous cyst with scolex | Retina | NA | Praziquantel and corticosteroids | NA | NA | Drug failed to destroy cyst |
| Topilow et al 1981 | USA | 47/M | 1 week | NA | Vision loss right  20/70  Left 20/25 | NA | A subretinal Intravetreous cyst with scolex both side  Retinal detachment | Retina | Cysticercal larva with scolex | NA | NA | Excision of the cyst | Vision not improved much |
| Zinn et al 1980 | USA | 13/F | 1 week | Seizures | Vision loss  Left  20/50  Right 20/30 | NA | Two cysts with scolex attached with optic nerve head on left side  Retinal detachment | Optic nerve | Cysticercal larva with scolex | Corticosteroids | NA | Excision of the cyst | Vision stable |
| Friedman et al 1980 | USA | 13/F | NA | Seizures | Vision loss  Left  20/50  Right 20/30 | NA | Two cysts with scolex in left vitreous cavity | Retina | Cysticercal larva with scolex | NA | NA | Excision of the cyst | Vision stable |
| Messner and Kammerer 1979 | USA | 59/F | NA | NA | Vision loss right 20/100 | NA | Two retinal cysts with scolex | Retina | Cysticercal larva with scolex | NA | NA | Excision of the cyst | Vision stable |
| Jain et al 1979 | India | 45/M | 4 years | NA | Vision loss left | NA | A cyst with scolex in left vitreous cavity | Retina | Cysticercal larva with scolex | NA | NA | Eye was removed | NA |
|  |  | 30/M | 4 years | NA | Vision loss left | NA | A cyst with scolex in retina  Retinal detachment | Retina | NA | NA | NA | NA | NA |
|  |  | 25/M | 2 years | Seizures  Subcutaneous nodules | Vision loss left finger counting only  papilloedema | NA | A cyst with scolex in retina | Retina | Subcutaneous nodule = Cysticercal larva with scolex | NA | NA | NA | NA |
|  |  | 32/M | 3 months | NA | Vision loss right | NA | A cyst with scolex in retina | Retina | NA | NA | NA | NA | NA |
|  |  | 22/M | 25 days | NA | Vision loss right | NA | A cyst with scolex in retina in macular area | Retina | NA | NA | NA | NA | NA |
|  |  | 11/M | 7days | NA | Vision loss right  6/24 | NA | A cyst with scolex in retina in macular area | Retina | NA | NA | NA | NA | NA |
|  |  | 26/F | 4 months | Headache  Seizures  Subcutaneous nodules | Vision loss bilateral  Papilledema  Proptosis | NA | A cyst with scolex in retina in left side macular area | Retina | Subcutaneous nodule = Cysticercal larva with scolex | NA | NA | NA | NA |
|  |  | 20/M | 7 days | NA | Vision loss left | NA | A cyst with scolex in retina in macular area | Retina | NA | NA | NA | NA | NA |
|  |  | 16/F | 1 year | NA | Vision loss left | NA | A cyst with scolex in retina in macular area | Retina | NA | NA | NA | NA | NA |
|  |  | 16/M | NA | Cranial nerve palsy  Hemiplegia | Vision loss bilateral 6/9   bilateral papilloedema | NA | A cyst with scolex in retina in right side | Retina | NA | NA | NA | NA | NA |
| Kapoor et al 1977 | India | 14/M | 2 weeks | NA | Vision loss right finger counting only | NA | A floating cyst with scolex in anterior chamber | Anterior chamber | Cysticercal larva with scolex | NA | NA | Excision of the cyst | Vision improved |
| Hutton et al 1976 | USA | 23/M | 10  days | NA | Vision loss right 20/60 | NA | Intravetreous  cyst | Retina | Cysticercal larva with scolex | NA | NA | Excision of the cyst | Vision improved |
| Bartholomew  1975 | UK | 25/F | NA | NA | Vision loss right finger counting  Right disk edema | NA | A subretinal cyst | Retina | Cysticercal larva with scolex | NA | NA | Excision of the cyst | Severe vitreous hemorrhage  Vision lost |
|  |  | 20/M | NA | NA | Vision loss right  20/200 | NA | A subretinal cyst  Retinal hemorrhage | Retina | NA | NA | NA | NA | NA |
| Manschot  1968 | Netherlands | 16/F | NA | NA | Vision loss right | NA | A subretinal cyst | Retina | Cysticercal larva with scolex | NA | NA | Removal of whole eye | NA |
| Dunlap 1965 | USA | 39/M | 2 years | NA | Vision loss left limited to light perception | NA | An Intravetreous cyst  Retinal detachment | Retina | Cysticercal larva with scolex | NA | NA | Excision of the cyst | No change in vision |
| Segal et al 1964 | Poland | 23/M | NA | NA | Vision loss right finger counting | NA | An Intravetreous cyst  Retinal detachment and retinal hemorrhage | Retina | Cysticercal larva with scolex | NA | NA | Excision of the cyst | Vision improved |
