## Supplementary item-2 for "Vision Loss in Neurocysticercosis: Clinical Features, Diagnostic Approaches, and Treatment Outcomes: a systematic review of case report and case series"

**Supplementary table-2: Evaluation of the methodological quality of case reports and case series**

| **Reference** | **Does the patient represent the whole experience of the investigator** | **Was the exposure adequately ascertained?** | **Was the outcome adequately ascertained?** | **Were other alternative causes that may explain the observation ruled out?** | **Was there a challenge and/or re-challenge phenomenon?** | **Was there a dose-response effect?** | **Was follow-up long enough for outcomes to occur?** | **Is the case(s) described with sufficient details to allow practitioners make inferences related to their own practice?** | **Score** |
| --- | --- | --- | --- | --- | --- | --- | --- | --- | --- |
| **Konar et al 2024** | **Yes** | **Yes** | **Yes** | **Yes** | **NA** | **Yes** | **Yes** | **Yes** | **7** |
| **Thangamathesvaran et al 2023** | **Yes** | **Yes** | **Yes** | **Yes** | **NA** | **Yes** | **No** | **Yes** | **6** |
| **Rehman et al 2023** | **Yes** | **Yes** | **Yes** | **Yes** | **NA** | **Yes** | **Yes** | **Yes** | **7** |
| **Rama et al 2023** | **Yes** | **Yes** | **Yes** | **Yes** | **NA** | **Yes** | **Yes** | **Yes** | **7** |
| **Mansour and Tahir 2023** | **Yes** | **Yes** | **Yes** | **Yes** | **NA** | **Yes** | **Yes** | **Yes** | **7** |
| **Grover et al 2023** | **Yes** | **Yes** | **Yes** | **Yes** | **NA** | **Yes** | **Yes** | **Yes** | **7** |
| **Gasca-Sánchez et al 2023** | **Yes** | **Yes** | **Yes** | **Yes** | **NA** | **Yes** | **Yes** | **Yes** | **7** |
| **Fan et al 2023** | **Yes** | **Yes** | **Yes** | **Yes** | **NA** | **Yes** | **Yes** | **Yes** | **7** |
|  | **Yes** | **Yes** | **Yes** | **Yes** | **NA** | **Yes** | **Yes** | **Yes** | **7** |
| **Das et al 2023** | **Yes** | **Yes** | **Yes** | **Yes** | **NA** | **Yes** | **Yes** | **Yes** | **7** |
| **Burgos-Sosa et al 2023** | **Yes** | **Yes** | **Yes** | **Yes** | **NA** | **Yes** | **Yes** | **Yes** | **7** |
|  | **Yes** | **Yes** | **Yes** | **Yes** | **NA** | **Yes** | **Yes** | **Yes** | **7** |
|  | **Yes** | **Yes** | **Yes** | **Yes** | **NA** | **Yes** | **No** | **Yes** | **6** |
| **Bansal et al 2023** | **Yes** | **Yes** | **Yes** | **Yes** | **NA** | **Yes** | **Yes** | **Yes** | **7** |
| **Koonwar et al 2022** | **Yes** | **Yes** | **Yes** | **Yes** | **NA** | **Yes** | **Yes** | **Yes** | **7** |
| **Opara 2022** | **Yes** | **Yes** | **Yes** | **Yes** | **NA** | **Yes** | **Yes** | **Yes** | **7** |
| **Jethva et al 2022** | **Yes** | **Yes** | **Yes** | **Yes** | **NA** | **Yes** | **NA** | **NA** | **5** |
| **Li et al 2022** | **Yes** | **Yes** | **Yes** | **Yes** | **NA** | **Yes** | **Yes** | **Yes** | **7** |
| **Koirala et al 2022** | **Yes** | **Yes** | **Yes** | **Yes** | **NA** | **Yes** | **Yes** | **Yes** | **7** |
| **Shrestha et al 2021** | **Yes** | **Yes** | **Yes** | **Yes** | **NA** | **Yes** | **Yes** | **Yes** | **7** |
| **Shakya et al 2021** | **Yes** | **Yes** | **Yes** | **Yes** | **NA** | **Yes** | **Yes** | **Yes** | **7** |
| **Samanta et al 2021** | **Yes** | **Yes** | **Yes** | **Yes** | **NA** | **Yes** | **Yes** | **Yes** | **7** |
| **Niang et al 2021** | **Yes** | **Yes** | **Yes** | **Yes** | **NA** | **Yes** | **NA** | **NA** | **5** |
| **Kumar and Padhy 2021** | **Yes** | **Yes** | **Yes** | **Yes** | **NA** | **Yes** | **Yes** | **Yes** | **7** |
| **Gala et al 2021** | **Yes** | **Yes** | **Yes** | **Yes** | **NA** | **Yes** | **NA** | **NA** | **5** |
| **Bhatia et al 2021** | **Yes** | **Yes** | **Yes** | **Yes** | **NA** | **Yes** | **Yes** | **Yes** | **7** |
| **Snyder et al 2020** | **Yes** | **Yes** | **Yes** | **Yes** | **NA** | **Yes** | **Yes** | **Yes** | **7** |
| **Silva et al 2020** | **Yes** | **Yes** | **Yes** | **Yes** | **NA** | **Yes** | **Yes** | **Yes** | **7** |
| **Shashni et al 2020** | **Yes** | **Yes** | **Yes** | **Yes** | **NA** | **Yes** | **NA** | **NA** | **5** |
| **Samanta et al 2020** | **Yes** | **Yes** | **Yes** | **Yes** | **NA** | **Yes** | **Yes** | **Yes** | **7** |
|  | **Yes** | **NA** | **Yes** | **Yes** | **NA** | **Yes** | **NA** | **NA** | **4** |
| **Reddy et al 2020** | **Yes** | **Yes** | **Yes** | **Yes** | **NA** | **Yes** | **NA** | **NA** | **5** |
| **Patidar et al 2020** | **Yes** | **Yes** | **Yes** | **Yes** | **NA** | **Yes** | **NA** | **Yes** | **6** |
| **Neto et al 2020** | **Yes** | **Yes** | **Yes** | **Yes** | **NA** | **Yes** | **Yes** | **Yes** | **7** |
| **Lee et al 2020** | **Yes** | **Yes** | **Yes** | **Yes** | **NA** | **Yes** | **Yes** | **Yes** | **7** |
| **Kumar et al 2020** | **Yes** | **Yes** | **Yes** | **Yes** | **NA** | **Yes** | **Yes** | **Yes** | **7** |
| **García Franco et al 2020** | **Yes** | **Yes** | **Yes** | **Yes** | **NA** | **Yes** | **Yes** | **Yes** | **7** |
| **Dhiman et al 2020** | **Yes** | **Yes** | **Yes** | **Yes** | **NA** | **Yes** | **Yes** | **Yes** | **7** |
| **Cuellar-Hernandez et al 2020** | **Yes** | **Yes** | **Yes** | **Yes** | **NA** | **Yes** | **Yes** | **Yes** | **7** |
| **Zou et al 2019** | **Yes** | **Yes** | **Yes** | **Yes** | **NA** | **Yes** | **Yes** | **Yes** | **7** |
|  | **Yes** | **Yes** | **Yes** | **Yes** | **NA** | **Yes** | **Yes** | **Yes** | **7** |
|  | **Yes** | **Yes** | **Yes** | **Yes** | **NA** | **Yes** | **Yes** | **Yes** | **7** |
| **Wadhwani et al 2019** | **Yes** | **Yes** | **Yes** | **Yes** | **NA** | **Yes** | **Yes** | **Yes** | **7** |
| **Shrestha and Shrestha 2019** | **Yes** | **Yes** | **Yes** | **Yes** | **NA** | **Yes** | **NA** | **Yes** | **6** |
| **Musara et al 2019** | **Yes** | **Yes** | **Yes** | **Yes** | **NA** | **Yes** | **Yes** | **Yes** | **7** |
| **Kumar et al 2019** | **Yes** | **Yes** | **Yes** | **Yes** | **NA** | **Yes** | **Yes** | **Yes** | **7** |
| **Chaugule et al 2019** | **Yes** | **Yes** | **Yes** | **Yes** | **NA** | **Yes** | **Yes** | **Yes** | **7** |
| **Swamy et al 2018** | **Yes** | **Yes** | **Yes** | **Yes** | **NA** | **Yes** | **Yes** | **Yes** | **7** |
| **Mukhija et al 2018** | **Yes** | **Yes** | **Yes** | **Yes** | **NA** | **Yes** | **Yes** | **Yes** | **7** |
| **Matalia et al 2018** | **Yes** | **Yes** | **Yes** | **Yes** | **NA** | **Yes** | **Yes** | **Yes** | **7** |
| **Goel 2018** | **Yes** | **Yes** | **Yes** | **Yes** | **NA** | **Yes** | **Yes** | **Yes** | **7** |
| **Obedulla et al 2017** | **Yes** | **Yes** | **Yes** | **Yes** | **NA** | **Yes** | **Yes** | **Yes** | **7** |
| **Majumdar and Pal 2017** | **Yes** | **NA** | **Yes** | **Yes** | **NA** | **Yes** | **NA** | **NA** | **4** |
| **Karthikeya et al 2017** | **Yes** | **Yes** | **Yes** | **Yes** | **NA** | **Yes** | **Yes** | **Yes** | **7** |
| **Gaur et al 2017** | **Yes** | **Yes** | **Yes** | **Yes** | **NA** | **Yes** | **Yes** | **Yes** | **7** |
| **Singh et al 2016** | **Yes** | **Yes** | **Yes** | **Yes** | **NA** | **Yes** | **Yes** | **Yes** | **7** |
| **Nie et al 2016** | **Yes** | **Yes** | **Yes** | **Yes** | **NA** | **Yes** | **NA** | **Yes** | **6** |
| **Krupa et al 2016** | **Yes** | **Yes** | **Yes** | **Yes** | **NA** | **Yes** | **Yes** | **Yes** | **7** |
| **Khalid et al 2016** | **Yes** | **Yes** | **Yes** | **Yes** | **NA** | **Yes** | **Yes** | **Yes** | **7** |
| **Jain et al 2016** | **Yes** | **Yes** | **Yes** | **Yes** | **NA** | **Yes** | **Yes** | **Yes** | **7** |
| **Wang and Huang 2015** | **Yes** | **Yes** | **Yes** | **Yes** | **NA** | **Yes** | **Yes** | **Yes** | **7** |
| **Vaitheeswaran et al 2015** | **Yes** | **Yes** | **Yes** | **Yes** | **NA** | **Yes** | **Yes** | **Yes** | **7** |
| **Singh and Singh**  **2015** | **Yes** | **Yes** | **Yes** | **Yes** | **NA** | **Yes** | **Yes** | **Yes** | **7** |
| **Sharma et al 2015** | **Yes** | **Yes** | **Yes** | **Yes** | **NA** | **Yes** | **Yes** | **Yes** | **7** |
| **Sharma et al 2015** | **Yes** | **Yes** | **Yes** | **Yes** | **NA** | **Yes** | **Yes** | **Yes** | **7** |
| **Nehasudhakar et al 2015** | **Yes** | **Yes** | **Yes** | **Yes** | **NA** | **Yes** | **NA** | **Yes** | **6** |
| **Narra et al 2015** | **Yes** | **Yes** | **Yes** | **Yes** | **NA** | **Yes** | **Yes** | **Yes** | **7** |
| **Mishra et al 2015** | **Yes** | **Yes** | **Yes** | **Yes** | **NA** | **Yes** | **Yes** | **Yes** | **7** |
| **Jain et al 2015** | **Yes** | **Yes** | **Yes** | **Yes** | **NA** | **Yes** | **Yes** | **Yes** | **7** |
| **Huang and Sridhar 2015** | **Yes** | **Yes** | **Yes** | **Yes** | **NA** | **Yes** | **Yes** | **Yes** | **7** |
| **Chavala et al 2015** | **Yes** | **Yes** | **Yes** | **Yes** | **NA** | **Yes** | **Yes** | **Yes** | **7** |
| **Chaudhary et al 2015** | **Yes** | **Yes** | **Yes** | **Yes** | **NA** | **Yes** | **Yes** | **Yes** | **7** |
| **Wani et al 2014** | **Yes** | **Yes** | **Yes** | **Yes** | **NA** | **Yes** | **Yes** | **Yes** | **7** |
| **Tran et al 2014** | **Yes** | **Yes** | **Yes** | **Yes** | **NA** | **Yes** | **Yes** | **Yes** | **7** |
| **Takkar et al 2014** | **Yes** | **Yes** | **Yes** | **Yes** | **NA** | **Yes** | **Yes** | **Yes** | **7** |
| **Sune et al 2014** | **Yes** | **Yes** | **Yes** | **Yes** | **NA** | **Yes** | **Yes** | **Yes** | **7** |
| **Salcedo-Villanueva et al 2014** | **Yes** | **Yes** | **Yes** | **Yes** | **NA** | **Yes** | **NA** | **Yes** | **6** |
| **Mohan et al 2014** | **Yes** | **Yes** | **Yes** | **Yes** | **NA** | **Yes** | **Yes** | **Yes** | **7** |
| **Matalia et al 2014** | **Yes** | **Yes** | **Yes** | **Yes** | **NA** | **Yes** | **Yes** | **Yes** | **7** |
| **Kasundra et al 2014** | **Yes** | **Yes** | **Yes** | **Yes** | **NA** | **Yes** | **Yes** | **Yes** | **7** |
| **Das et al 2014** | **Yes** | **Yes** | **Yes** | **Yes** | **NA** | **Yes** | **Yes** | **Yes** | **7** |
| **Ko et al 2013** | **Yes** | **Yes** | **Yes** | **Yes** | **NA** | **Yes** | **Yes** | **Yes** | **5** |
| **Kori et al 2013** | **Yes** | **NA** | **Yes** | **Yes** | **NA** | **Yes** | **NA** | **Yes** | **7** |
| **Babalola et al 2013** | **Yes** | **Yes** | **Yes** | **Yes** | **NA** | **NA** | **NA** | **NA** | **4** |
| **Aulakh 2013** | **Yes** | **Yes** | **Yes** | **Yes** | **NA** | **Yes** | **Yes** | **Yes** | **7** |
| **Venna et al 2012** | **Yes** | **Yes** | **Yes** | **Yes** | **NA** | **Yes** | **Yes** | **Yes** | **7** |
| **Swastika et al 2012** | **Yes** | **Yes** | **Yes** | **Yes** | **NA** | **Yes** | **Yes** | **Yes** | **7** |
| **Sinha S et al 2012** | **Yes** | **Yes** | **Yes** | **Yes** | **NA** | **Yes** | **NA** | **Yes** | **6** |
| **Raval et al 2012** | **Yes** | **Yes** | **Yes** | **Yes** | **NA** | **Yes** | **NA** | **Yes** | **6** |
| **Prasad et al 2012** | **Yes** | **Yes** | **Yes** | **Yes** | **NA** | **Yes** | **Yes** | **Yes** | **7** |
| **Parashari et al 2012** | **Yes** | **Yes** | **Yes** | **Yes** | **NA** | **Yes** | **Yes** | **Yes** | **5** |
| **Mulla et al 2012** | **Yes** | **Yes** | **Yes** | **Yes** | **NA** | **Yes** | **Yes** | **Yes** | **5** |
| **Hussain et al 2012** | **Yes** | **Yes** | **Yes** | **Yes** | **NA** | **Yes** | **Yes** | **Yes** | **7** |
| **Agarwal et al 2012** | **Yes** | **Yes** | **Yes** | **Yes** | **NA** | **Yes** | **Yes** | **Yes** | **7** |
| **Sundar et al 2011** | **Yes** | **Yes** | **Yes** | **Yes** | **NA** | **Yes** | **Yes** | **Yes** | **5** |
| **Ratra et al 2010** | **Yes** | **Yes** | **Yes** | **Yes** | **NA** | **Yes** | **Yes** | **Yes** | **7** |
|  | **Yes** | **Yes** | **Yes** | **Yes** | **NA** | **Yes** | **Yes** | **Yes** | **7** |
| **Prasad et al 2010** | **Yes** | **Yes** | **Yes** | **Yes** | **NA** | **Yes** | **Yes** | **Yes** | **7** |
| **Dass et al 2010** | **Yes** | **Yes** | **Yes** | **Yes** | **NA** | **Yes** | **Yes** | **Yes** | **7** |
| **Cheong et al 2010** | **Yes** | **Yes** | **Yes** | **Yes** | **NA** | **Yes** | **Yes** | **Yes** | **7** |
| **Yadav et al 2009** | **Yes** | **Yes** | **Yes** | **Yes** | **NA** | **Yes** | **Yes** | **Yes** | **7** |
|  | **Yes** | **Yes** | **Yes** | **Yes** | **NA** | **Yes** | **Yes** | **Yes** | **7** |
|  | **Yes** | **Yes** | **Yes** | **Yes** | **NA** | **Yes** | **Yes** | **Yes** | **7** |
| **Taksande et al 2009** | **Yes** | **Yes** | **Yes** | **Yes** | **NA** | **Yes** | **Yes** | **Yes** | **7** |
| **Patnaik et al 2009** | **Yes** | **Yes** | **Yes** | **Yes** | **NA** | **Yes** | **Yes** | **Yes** | **7** |
| **Miller et al 2009** | **Yes** | **Yes** | **Yes** | **Yes** | **NA** | **Yes** | **Yes** | **Yes** | **7** |
| **Knight et al 2009** | **Yes** | **Yes** | **Yes** | **Yes** | **NA** | **Yes** | **Yes** | **Yes** | **7** |
| **Venkatesh et al 2008** | **Yes** | **Yes** | **Yes** | **Yes** | **NA** | **Yes** | **Yes** | **Yes** | **7** |
| **Kai et al 2008** | **Yes** | **Yes** | **Yes** | **Yes** | **NA** | **Yes** | **Yes** | **Yes** | **7** |
| **Aghamohammadi et al 2008** | **Yes** | **Yes** | **Yes** | **Yes** | **NA** | **Yes** | **NA** | **Yes** | **6** |
| **Shariq and Adhikari 2007** | **Yes** | **Yes** | **Yes** | **Yes** | **NA** | **Yes** | **Yes** | **Yes** | **7** |
| **Mahendradas et al 2007** | **Yes** | **Yes** | **Yes** | **Yes** | **NA** | **Yes** | **Yes** | **Yes** | **7** |
| **Karande and Kumbhare 2007** | **Yes** | **Yes** | **Yes** | **Yes** | **NA** | **Yes** | **Yes** | **Yes** | **7** |
| **Goyal et al 2007** | **Yes** | **Yes** | **Yes** | **Yes** | **NA** | **Yes** | **Yes** | **Yes** | **7** |
| **Cortez et al 2007** | **Yes** | **Yes** | **Yes** | **Yes** | **NA** | **Yes** | **Yes** | **Yes** | **7** |
| **Sudan et al 2005** | **Yes** | **Yes** | **Yes** | **Yes** | **NA** | **Yes** | **Yes** | **Yes** | **7** |
| **Pushker et al 2005** | **Yes** | **Yes** | **Yes** | **Yes** | **NA** | **Yes** | **Yes** | **Yes** | **7** |
| **Chadha et al 2005** | **Yes** | **Yes** | **Yes** | **Yes** | **NA** | **Yes** | **Yes** | **Yes** | **7** |
| **Besada et al 2005** | **Yes** | **Yes** | **Yes** | **Yes** | **NA** | **Yes** | **Yes** | **Yes** | **7** |
| **Lim and Chee 2004** | **Yes** | **Yes** | **Yes** | **Yes** | **NA** | **Yes** | **Yes** | **Yes** | **7** |
| **Bowie et al 2004** | **Yes** | **Yes** | **Yes** | **Yes** | **NA** | **Yes** | **Yes** | **Yes** | **7** |
| **Hiralal et al 2003** | **Yes** | **Yes** | **Yes** | **Yes** | **NA** | **Yes** | **Yes** | **Yes** | **7** |
| **Adegbehingbe et al 2003** | **Yes** | **Yes** | **Yes** | **Yes** | **NA** | **NA** | **NA** | **NA** | **4** |
| **Verma et al 2002** | **Yes** | **Yes** | **Yes** | **Yes** | **NA** | **Yes** | **Yes** | **Yes** | **7** |
| **Das et al 2002** | **Yes** | **Yes** | **Yes** | **Yes** | **NA** | **Yes** | **Yes** | **Yes** | **7** |
| **Chung et al 2002** | **Yes** | **Yes** | **Yes** | **Yes** | **NA** | **Yes** | **Yes** | **Yes** | **5** |
| **Bajaj and Pushker 2002** | **Yes** | **Yes** | **Yes** | **Yes** | **NA** | **Yes** | **Yes** | **Yes** | **7** |
| **Sidhu et al 2001** | **Yes** | **Yes** | **NA** | **Yes** | **NA** | **NA** | **NA** | **NA** | **3** |
| **Sabti et al 2001** | **Yes** | **Yes** | **Yes** | **Yes** | **NA** | **Yes** | **Yes** | **Yes** | **7** |
| **Lombardo 2001** | **Yes** | **Yes** | **Yes** | **Yes** | **NA** | **Yes** | **Yes** | **Yes** | **7** |
| **Chandra et al 2000** | **Yes** | **Yes** | **Yes** | **Yes** | **NA** | **Yes** | **Yes** | **Yes** | **7** |
| **Gurha et al 1999** | **Yes** | **Yes** | **Yes** | **Yes** | **NA** | **Yes** | **Yes** | **Yes** | **7** |
| **Betharia et al 1999** | **Yes** | **Yes** | **Yes** | **Yes** | **NA** | **Yes** | **Yes** | **Yes** | **7** |
| **Tandon et al 1998** | **Yes** | **Yes** | **Yes** | **Yes** | **NA** | **Yes** | **Yes** | **Yes** | **7** |
|  | **Yes** | **Yes** | **Yes** | **Yes** | **NA** | **Yes** | **Yes** | **Yes** | **7** |
| **Gupta et al 1998** | **Yes** | **Yes** | **Yes** | **Yes** | **NA** | **Yes** | **Yes** | **Yes** | **7** |
| **George et al 1998** | **Yes** | **Yes** | **Yes** | **Yes** | **NA** | **Yes** | **Yes** | **Yes** | **7** |
| **Seo et al 1996** | **Yes** | **Yes** | **Yes** | **Yes** | **NA** | **Yes** | **Yes** | **Yes** | **7** |
| **Bousquet et al 1996** | **Yes** | **Yes** | **Yes** | **Yes** | **NA** | **Yes** | **Yes** | **Yes** | **7** |
| **Santoyo et al 1991** | **Yes** | **Yes** | **Yes** | **Yes** | **NA** | **Yes** | **Yes** | **Yes** | **7** |
| **Mason et al 1991** | **Yes** | **Yes** | **Yes** | **Yes** | **NA** | **Yes** | **Yes** | **Yes** | **7** |
| **Luger et al 1991** | **Yes** | **Yes** | **Yes** | **Yes** | **NA** | **Yes** | **Yes** | **Yes** | **7** |
| **Steinmetz et al 1989** | **Yes** | **Yes** | **Yes** | **Yes** | **NA** | **Yes** | **Yes** | **Yes** | **7** |
| **Pansey et al 1989** | **Yes** | **Yes** | **Yes** | **Yes** | **NA** | **Yes** | **Yes** | **Yes** | **7** |
| **Rafael and Gomez-Llata 1985** | **Yes** | **Yes** | **Yes** | **Yes** | **NA** | **Yes** | **Yes** | **Yes** | **7** |
| **Kruger-Leite et al 1985** | **Yes** | **Yes** | **Yes** | **Yes** | **NA** | **Yes** | **Yes** | **Yes** | **7** |
| **Kestelyn and Taelman 1985** | **Yes** | **Yes** | **Yes** | **Yes** | **NA** | **Yes** | **Yes** | **Yes** | **7** |
| **Topilow et al 1981** | **Yes** | **Yes** | **Yes** | **Yes** | **NA** | **Yes** | **Yes** | **Yes** | **7** |
| **Zinn et al 1980** | **Yes** | **Yes** | **Yes** | **Yes** | **NA** | **Yes** | **Yes** | **Yes** | **7** |
| **Friedman et al 1980** | **Yes** | **Yes** | **Yes** | **Yes** | **NA** | **Yes** | **Yes** | **Yes** | **7** |
| **Messner and Kammerer 1979** | **Yes** | **Yes** | **Yes** | **Yes** | **NA** | **Yes** | **Yes** | **Yes** | **7** |
| **Jain et al 1979** | **Yes** | **Yes** | **NA** | **Yes** | **NA** | **NA** | **NA** | **NA** | **3** |
|  | **Yes** | **Yes** | **NA** | **Yes** | **NA** | **NA** | **NA** | **NA** | **3** |
|  | **Yes** | **Yes** | **NA** | **Yes** | **NA** | **NA** | **NA** | **NA** | **3** |
|  | **Yes** | **Yes** | **NA** | **Yes** | **NA** | **NA** | **NA** | **NA** | **3** |
|  | **Yes** | **Yes** | **NA** | **Yes** | **NA** | **NA** | **NA** | **NA** | **3** |
|  | **Yes** | **Yes** | **NA** | **Yes** | **NA** | **NA** | **NA** | **NA** | **3** |
|  | **Yes** | **Yes** | **NA** | **Yes** | **NA** | **NA** | **NA** | **NA** | **3** |
|  | **Yes** | **Yes** | **NA** | **Yes** | **NA** | **NA** | **NA** | **NA** | **3** |
|  | **Yes** | **Yes** | **NA** | **Yes** | **NA** | **NA** | **NA** | **NA** | **3** |
|  | **Yes** | **Yes** | **NA** | **Yes** | **NA** | **NA** | **NA** | **NA** | **3** |
| **Kapoor et al 1977** | **Yes** | **Yes** | **Yes** | **Yes** | **NA** | **Yes** | **Yes** | **Yes** | **7** |
| **Hutton et al 1976** | **Yes** | **Yes** | **Yes** | **Yes** | **NA** | **Yes** | **Yes** | **Yes** | **7** |
| **Bartholomew**  **1975** | **Yes** | **Yes** | **Yes** | **Yes** | **NA** | **Yes** | **Yes** | **Yes** | **7** |
|  | **Yes** | **Yes** | **NA** | **Yes** | **NA** | **NA** | **NA** | **NA** | **3** |
| **Manschot**  **1968** | **Yes** | **Yes** | **Yes** | **Yes** | **NA** | **Yes** | **Yes** | **Yes** | **7** |
| **Dunlap 1965** | **Yes** | **Yes** | **Yes** | **Yes** | **NA** | **Yes** | **Yes** | **Yes** | **7** |
| **Segal et al 1964** | **Yes** | **Yes** | **Yes** | **Yes** | **NA** | **Yes** | **Yes** | **Yes** | **7** |

**Total= 168**

**Score 6, 7 = 142 (85%)**

**Score less than 5 or less than 5 = 26 (15%)**

**Domains Leading explanatory questions**

Selection 1. Does the patient(s) represent(s) the whole experience of the investigator or is the selection method unclear to the extent that other patients with similar presentation may not have been reported?

Ascertainment 2. Was the exposure adequately ascertained?

3. Was the outcome adequately ascertained?

Causality

4. Were other alternative causes that may explain the observation ruled out?

5. Was there a challenge/re-challenge phenomenon?

6. Was there a dose–response effect?

7. Was follow-up long enough for outcomes to occur?

Reporting

8. Is the case(s) described with sufficient details to allow other investigators to replicate the research or to allow practitioners make

inferences related to their own practice?
